# Cell-free DNA Fragmentomics Assay to Discriminate the Malignancy of Breast Nodules and Evaluate Treatment Response

**DOI:** 10.1101/2024.10.15.24315518

**Authors:** Jiaqi Liu, Yalun Li, Wanxiangfu Tang, Lijun Dai, Ziqi Jia, Heng Cao, Chenghao Li, Yuchen Liu, Yansong Huang, Jiang Wu, Dongxu Ma, Guangdong Qiao, Hua Bao, Shuang Chang, Dongqin Zhu, Shanshan Yang, Xuxiaochen Wu, Xue Wu, Hengyi Xu, Hongyan Chen, Yang Shao, Xiang Wang, Zhihua Liu, Jianzhong Su

**Author notes:** Corresponding authors. (Su J); (Liu Z); (Wang X). These authors contributed equally to this article (Jiaqi Liu, Yalun Li, Wanxiangfu Tang, Lijun Dai, Ziqi Jia).

## Abstract

The fragmentomics-based cell-free DNA (cfDNA) assays have recently illustrated prominent abilities to identify various cancers from non-conditional healthy controls, while their accuracy for identifying early-stage cancers from benign lesions with inconclusive imaging results remains uncertain. Especially for breast cancer, current imaging-based screening methods suffer from high false-positive rates for women with breast nodules, leading to unnecessary biopsies, which add to discomfort and healthcare burden. Here, we enroll 560 female participants in this multi-center study and demonstrate that cfDNA fragmentomics is a robust non-invasive biomarker for breast cancer using whole-genome sequencing. Among the multimodal cfDNA fragmentomics profiles, the fragment size ratio (FSR), fragment size distribution (FSD), and copy number variation (CNV) show more distinguishing ability than Griffin, motif breakpoint (MBP), and neomer. The cfDNA fragmentomics (cfFrag) model using the optimal three fragmentomics features discriminated early-stage breast cancers from benign nodules, even at a low sequencing depth (3×). Notably, it demonstrated a specificity of 94.1% in asymptomatic healthy women at a 90% sensitivity for breast cancers. Moreover, we comprehensively showcase the clinical utilities of the cfFrag model in predicting patient responses to neoadjuvant chemotherapy (NAC) and in combining with multimodal features, including radiological results and cfDNA methylation features (with AUC values of 0.93 – 0.94 and 0.96, respectively).

## Introduction

Breast cancer is one of the most common types of cancer worldwide and accounts for the highest number of cancer-related deaths among females [1]. Early detection of breast cancer is crucial for improving patients’ outcomes and survival [2]. However, current imaging-based screening methodologies, including mammography and ultrasonography, suffer from high false-positive rates, leading to many unnecessary biopsies, adding to patient discomfort [3]. Meanwhile, tumor biomarkers such as CA15-3 lack sensitivity for early-stage breast cancer [4]. Thus, liquid biopsies are needed as a non-invasive alternative or adjunct to select the high breast cancer-risk women for tumor biopsies [5].

Mutation-based circulating tumor DNA (ctDNA) detection has become the companion diagnostic by identifying actionable targets and alterations mediating resistance (e.g., *ESR1* and *PIK3CA* mutations in breast cancer) [6–8]. However, ctDNA typically lacks mutations, especially in early-stage disease, which limits its application in these contexts and reduces its ability to anticipate the diagnosis of localized cancer [9]. Besides, lacking common mutations in breast cancer limits the detection sensitivity in the patient-naïve approach [10]. Epigenetic analysis approaches offer potential solutions to fully exploit liquid biopsy in various settings [11, 12]. We previously conducted a whole-genome DNA methylation analysis on cell-free DNA (cfDNA) and identified ten optimal DNA methylation markers associated with breast cancer, which could enhance early detection [4]. However, current bisulfite-based methylation sequencing is prone to cfDNA damage, resulting in high cfDNA amount, depth dependencies, and increased cost.

Fragmentomics-based cfDNA assays have recently illustrated prominent abilities to identify various cancer types from paired non-conditional healthy controls using whole-genome sequencing (WGS) [13–17] and targeted cfDNA panels [18]. Similar to most cancer types, benign tumors also release ctDNA with unique features [19]. However, the accuracies of the cfDNA fragmentomics profile for identifying early-stage cancers from benign lesions with similar symptoms or inconclusive imaging results and predicting the therapeutic response remain largely unclear.

Herein, we developed a non-invasive liquid biopsy assay for early-stage breast cancer diagnosis which analyzes cfDNA fragmentomics through low-depth WGS and machine learning (**Figure 1**). To reveal its clinical utilities, we comprehensively evaluated the performances of this cfDNA fragmentomics assay in diagnosing early-stage breast cancer from benign breast nodules, predicting patient responses to neoadjuvant chemotherapy (NAC), and combining with multimodal features, including standard imaging techniques and cfDNA methylation markers. This approach is particularly beneficial for female patients who have undergone unnecessary biopsies due to false positives from imaging tests on benign breast nodules. Additionally, it can offer valuable insights into neoadjuvant treatment planning for breast cancer patients. Combining a cfDNA fragmentomics assay with standard imaging techniques enhances the early detection rate of breast cancer, potentially improving breast cancer survival rates.

**Figure 1.**
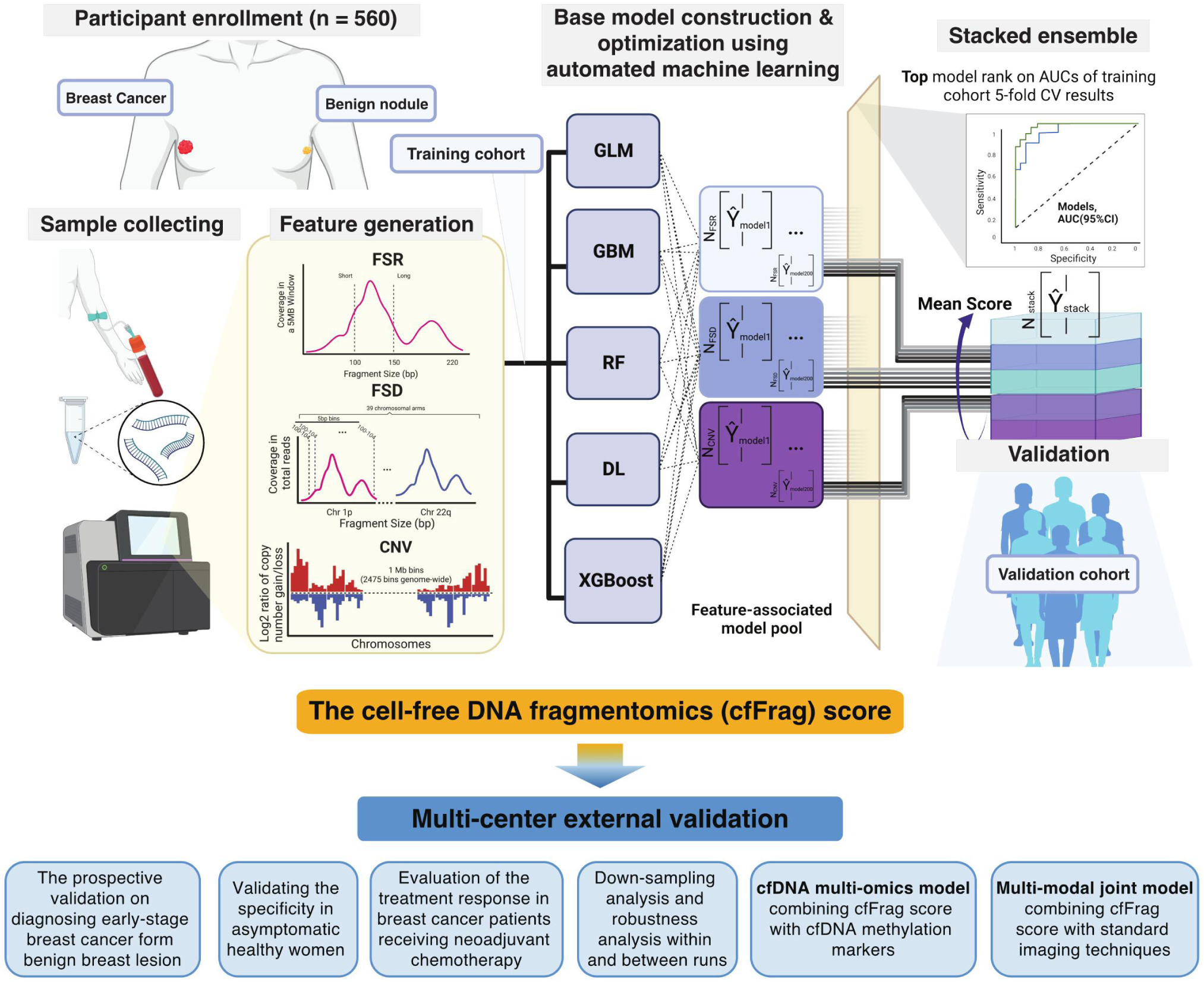
Study Design. Plasma samples collected from patients with breast cancers or benign nodules were used to extract cfDNA. WGS was performed on the cfDNA to generate fragmentomics feature types. The fragmentomics machine learning model was constructed using the optimal three fragmentomics profiles in the training set, including FSR, FSD, and CNV. Five different algorithms were utilized in the automatic machine-learning process. For each feature type, the top three models with the highest AUCs of 5-fold cross-validation in the training cohort were selected, and the mean cancer score (the cfFrag score) was used for the fragmentomics model. The cfFrag model was developed in the training cohort and evaluated in the validation cohorts. Abbreviation: cfDNA, cell-free DNA; WGS, whole-genome sequencing; FSR, fragment size ratio; FSD, fragment size distribution; CNV, copy number variation; GLM, generalized linear model; GBM, gradient boosting machine; RF, random forest; DL, deep learning; XGBoost, eXtreme gradient boosting; AUC, area under the curve; CV, cross-validation.

## Results

### Patient characteristics in two independent cohorts

We enrolled a total of 560 female participants in this multi-center study. In the training set, we enrolled 91 patients with breast cancers and 102 women with breast benign nodules from the Affiliated Yantai Yuhuangding Hospital of Qingdao University (YYH) in Yantai, China. Seven patients who refused to biopsy were excluded. In the external validation cohort, we recruited 143 patients with breast cancers and 66 women with benign nodules from the Cancer Hospital of the Chinese Academy of Medical Sciences (CHCAMS) in Beijing, China. The external screening cohort recruited 119 asymptomatic healthy women from our previous cohort of non-cancer healthy volunteers in Nanjing, China (Nanjing Cohort [14]). NAC validation cohort included 9/33 (27.3%) patients with pathological complete response (pCR) and 24/33 (72.7%) patients with non-pCR from the CHCAMS. The robustness analysis cohort contained three stage-II breast cancer patients and three patients with benign nodules (**Figure 2**).

**Figure 2.**
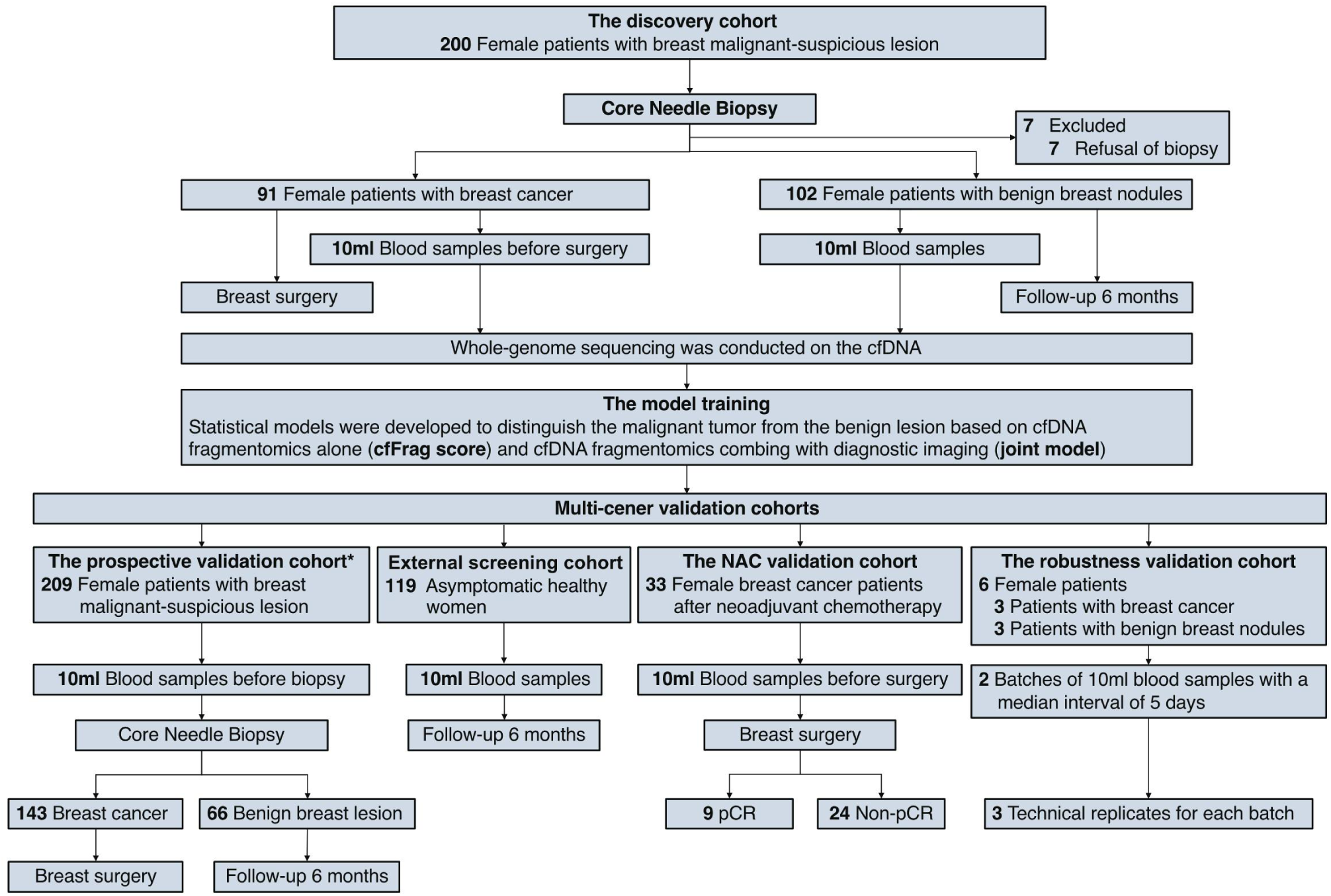
Patient Enrollment. In this multi-center study, we recruited 200 consecutive female patients with malignant-suspicious breast imaging results from the Yantai cohort as the training set. As a result, 91 patients with breast cancer and 102 women with benign breast nodules were enrolled, and seven patients who refused to biopsy were excluded. The external validation cohorts were composed of a screening cohort of healthy women in Nanjing (N = 119) and three independent validation cohorts in Beijing, namely the prospective validation cohort (N = 209), the neoadjuvant chemotherapy validation cohort (N = 33), and the robustness analysis cohort (N = 6). *The prospective validation cohort included 39 participants (14 with breast cancer and 25 with benign nodules) enrolled in a methylation-based early detection analysis for breast cancer through whole-genome bisulfite sequencing [4]. Abbreviation: NAC, neoadjuvant chemotherapy; pCR, pathological complete response.

The breast cancer patients enrolled in the training and validation cohorts were all in the early stages (0-II), including 8.8% and 16.0% in ductal carcinoma *in situ* (DCIS)/stage 0, 36.3% and 39.9% in stage I, 54.9% and 42.0% in stage II (Table S1). Among these patients, the majority type of breast cancer (80.4% – 85.7%) was invasive ductal carcinoma (IDC), and 16.1% – 18.7% of them were identified as triple-negative breast cancer (TNBC) in both cohorts.

#### Whole-genome multi-features analysis of cell-free DNA identifies the optimizing cfDNA fragmentomics profiles for breast cancer detection

In the Yantai cohort (training set), an average amount of 5.6 ng cfDNA (2.3 – 26.5 ng) was extracted from 500ul plasma. In the Beijing cohorts (validation set), 2 ml plasma was used to extract cfDNA for an average amount of 8.8 ng (3.4 – 13.5 ng). We applied low-depth WGS to the cfDNA samples. Libraries were sequenced in 7.4× mean depth (2.9 – 11.3×) in the training set and 8.8× mean depth (3.4 – 13.5×) in the validation set, resulting in a highly unique mapping rate and unique deduplicated mapping rate of more than 99.96%. The fragmentomics profiles were generated using low-depth WGS data from plasma cfDNA. To find optimal features for model construction, six types of fragmentomics profiles, including copy number variation (CNV), fragment size distribution (FSD), fragment size ratio (FSR), Griffin, motif breakpoint (MBP), and neomer, were generated using in-house scripts as previously reported [13, 15, 20–23]. Distinct spectrums of cfDNA fragmentomics features were found in patients with breast cancers and benign nodules, especially in CNV, FSD, and FSR (Figure S1).

We used the ichorCNA [15] reported tumor fraction (TF) to show the differences in CNV profile between breast cancer patients and benign nodule patients. As shown in Figure S2, the ichorCNA reported TF was significantly higher for the breast cancer patients than the benign nodule patients in both the training cohort (*P* = 8.0×10^−5^) and the validation cohort (*P* = 0.029). This suggests that while the breast cancer and benign groups both vary substantially from health baselines, there are still distinguishable differences between the two groups.

Next, base learners were constructed and optimized utilizing the machine-learning process utilizing five different algorithms of the machine-learning process [13] on the training set (Figure 1). Among the six fragmentomics features, CNV, FSD, and FSR demonstrated significantly higher area under the curve (AUC) values compared to all features (student’s *t*-test, *P* = 1.1×10^−3^, 4.3×10^−3^, and 2.7×10^−2^, respectively; **Figure 3A**).

**Figure 3.**
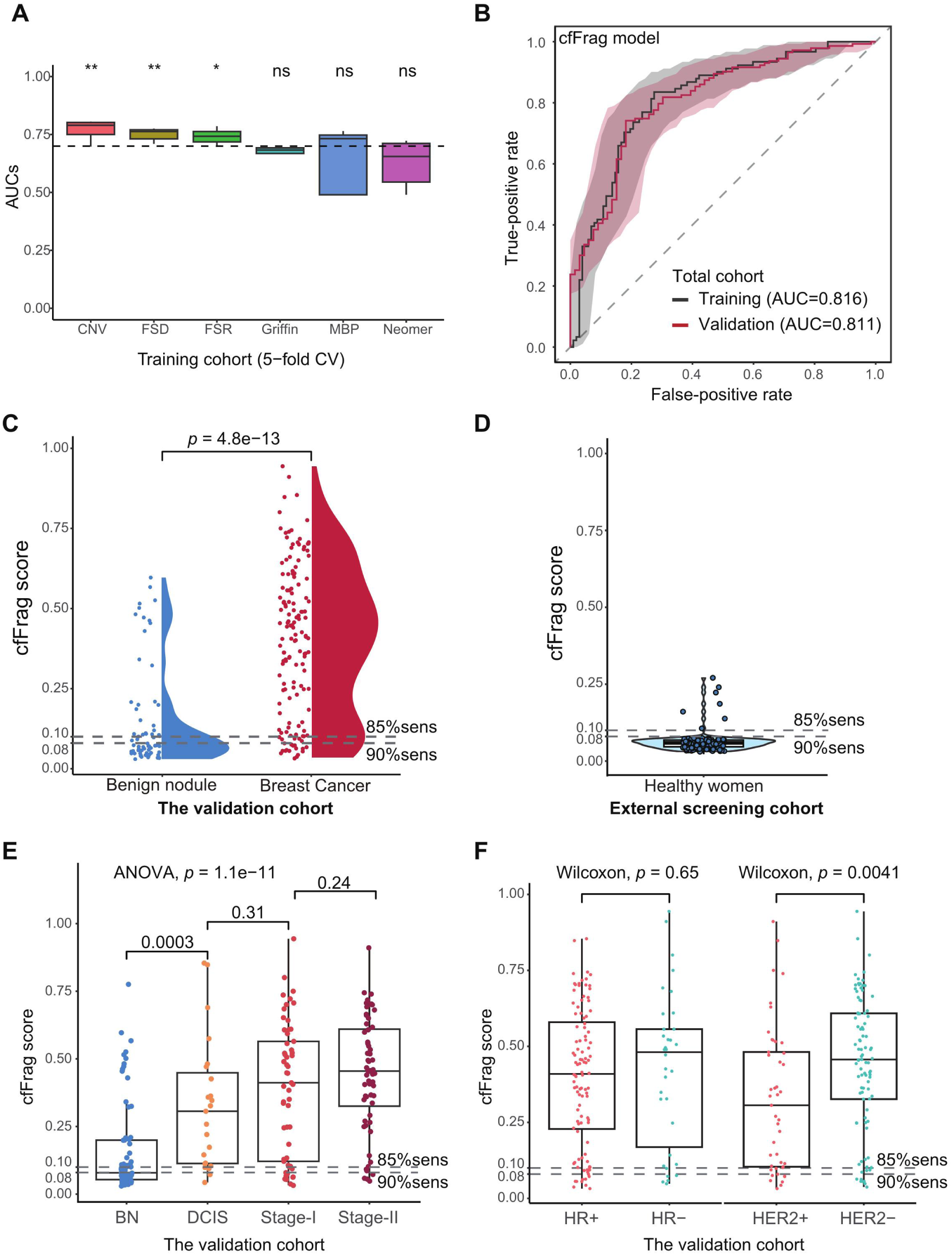
Evaluation of the cfDNA Fragmentomics (cfFrag) Model. **A.** Boxplots for AUCs of top base learners for six different cfDNA fragmentomics features. The t-test *P* values are 0.0011, 0.0043, 0.027, 0.17, 0.34, and 0.086 for these features, respectively. **B.** ROC curves using the training cohort (5-fold cross-validation) and the independent validation cohort. **C.** Violin plot illustrating cfFrag score distribution in the independent validation cohort’s benign nodule and breast cancer groups. The cutoffs, shown as the dotted lines, were determined by the training cohort. **D.** Violin plot using cfFrag cancer risk scores in 119 healthy female volunteers from our previous study [13]. The cfFrag scores for most healthy women (112/119) were lower than both cut-off values, yielding an excellent specificity of 94.1%. **E.** The box plot illustrating cfFrag score distribution in the benign nodule group and very early (DCIS), and early-stage (stages I and II) breast cancer groups. **F.** The box plot illustrating the cfFrag score distribution of different subgroups in the validation cohort. Abbreviation: CNV, copy number variation. FSD, fragment size distribution. FSR, fragment size ratio. MBP, motif breakpoint; AUC, area under the curve; CV, cross-validation; ns, not significant; sens, sensitivity; BN, benign nodule; ROC, receiver operating characteristic; DCIS, ductal carcinoma *in situ*; HR, hormone receptor; HER2, human epidermal growth factor receptor 2.

#### The cfDNA fragmentomics (cfFrag) model accurately distinguishes early-stage breast cancers from benign nodules with high specificity in asymptomatic healthy women

The cfDNA fragmentomics (cfFrag) scores were constructed using the optimal three fragmentomics profiles (CNV, FSR, and FSD) to predict breast cancers in the training cohort. A total of 24 (3 × 8) top base learners were selected to create the final cfFrag score by the 5-fold cross-validation AUC in the training cohort (Figures S3 and S4). Among the three feature types, CNV showed the highest mean AUC of 0.742 [0.661 – 0.791] for its top 8 base learners, while the FSD and FSR showed similar predict power in mean AUC (0.706 [0.631 – 0.750] and 0.706 [0.647 – 0.754]; Table S2). The top-performing features for each feature type were identified by summarizing the ranking of their relative importance in each base learner, as shown in Tables S3, S4, and S5, respectively.

To illustrate the impact of these top-performing features on the final cfFrag model, a recursive feature elimination analysis was performed. We constructed multiple cfFrag models using various subsets of top-performing features and evaluated their performances in the training and validation cohorts. The cfFrag model showed possible overfitting in the training cohort by using only subsets of top-performing features in the final model (Figure S5). The 5-fold cross-validation AUCs in the training cohort gradually decreased as more features were used in the model construction process. The fragmentomics model illustrated a solid discriminatory power between the breast cancer and benign nodules, yielding AUCs of 0.82 (95% CI: 0.76 – 0.88) and 0.81 (95% CI: 0.75 – 0.87) in training and external validation cohorts, respectively (Figure 3B).

The distribution patterns of the cfFrag scores showed significant differences between the benign nodule and breast cancer groups in the training cohort (Wilcoxon, *P* = 3.9×10^−14^; Figure S6), suggesting the cfFrag scores were positively associated with the probability of breast cancer. A similar trend was observed in the prospective validation cohort, with the breast cancer group showing a significantly higher cfFrag score than the benign nodule group (*P* = 4.8×10^−13^; Figure 3C). It achieved a specificity of 51.5% (95% CI: 38.9-64.0%) at the designed 90% sensitivity (95% CI: 83.3 – 94%) in the independent validation cohort, resulting in an overall accuracy of 77.5% (95% CI: 71.2 – 83.0%; **Table 1**). Setting the cut-off value at 85% sensitivity, the fragmentomics model reached specificities of 65.7% (95% CI: 55.6 – 74.8%) and 60.6% (95% CI: 47.8 – 72.4%) in the training and validation cohorts, respectively (Table S6).

**Table 1.** Evaluating the cfFrag Model Performances in the Training and Validation Cohorts.

|  | Training cohort<br>(5-fold cross-validation) | Prospective validation cohort | Asymptomatic healthy women |
| --- | --- | --- | --- |
| Breast cancer patients (n) | 91 | 143 | - |
| Benign nodule patients (n) | 102 | 66 | - |
| Healthy controls (n) | - | - | 119 |
| Sensitivity (95% CI) | 90.1% (82.1-95.4%) | 89.5% (83.3-94%) | - |
| Specificity (95% CI) | 52.0% (41.8-62.0%) | 51.5% (38.9-64%) | 94.1% (88.3-97.6%) |
| PPV (95% CI) | 62.6% (53.7-70.9%) | 80.0% (73.0-85.9%) | - |
| NPV (95% CI) | 85.5% (74.2-93.1%) | 69.4% (54.6-81.7%) | - |
| Accuracy (95% CI) | 69.9% (62.9-76.3%) | 77.5% (71.2-83%) | - |

Abbreviation: CI, confidence interval; PPV, positive predictive value; NPV, negative predictive value.

To verify the specificity of the cfFrag score in healthy women, we analyzed the cfDNA WGS data from 119 asymptomatic healthy women to generate the cfFrag scores. As a result, it yielded an excellent specificity of 94.1% (112/119, 95% CI: 88.3 – 97.6%; Table 1 and Figure 3D) for both cut-off values for 85% and 90% sensitivities.

#### The cfDNA fragmentomics (cfFrag) model maintains excellent performances in subgroup analysis and correlation with clinical features

To address the potential bias brought by the imbalanced age between breast cancer and benign nodule patients, a propensity score matching analysis was performed. We selected 112 patients (64 breast cancers and 48 benign nodules with matching ages) from the training cohort and 174 patients (117 breast cancers and 57 benign nodules with matching ages) from the prospective validation cohort. As a result, the fragmentomics model showed equally excellent predictive ability for breast cancer in these age-matched subsets, yielding AUCs of 0.82 (95% CI: 0.73 – 0.90) and 0.82 (95% CI: 0.75 – 0.88) in the training and validation cohort, respectively (Figure S7A). Similarly, the predictive model was able to maintain its predictive ability in a cohort containing small nodules (size ≤ 1cm), showing a high AUC of 0.83 (95% CI: 0.72 – 0.95) in the validation cohort (Figure S7B), compared to the traditional imaging methods (AUCs = 0.64 and 0.80 for mammography and ultrasound, respectively; Figure S8).

A subgroup analysis focused on the model’s performance was also performed to investigate potential bias. The sensitivities remained high for detecting various breast cancer subgroups, including the nodule size, stages, histology, and hormone receptor (HR) status (Figure S9). As the size of the benign nodules increased, the ability to identify different subgroups with specificity decreased (< 2cm: 54.9%, 2 – 5cm: 40.0%; Figure S10).

In addition to conducting subgroup analysis within the validation cohort, we performed a bootstrap analysis to minimize potential bias. Sensitivities derived from 100 bootstrap iterations for various breast cancer subgroups displayed patterns similar to our previous observations (Figure S11). Additionally, the specificities for the benign nodule subgroup, assessed through 100 bootstrap iterations, align with the trends seen in the validation cohort (Figure S12).

To demonstrate the performance for early detection, the cfFrag scores showed a significant gradual increase from the benign nodule to the DCIS and early-stage (stages I and II) breast cancer (ANOVA, *P* = 1.1×10^−11^; Figure 3E). Although the cfFrag score distribution showed no significant difference between the HR-positive and HR-negative groups, as well as between the TNBC and non-TNBC groups, the HER2-negative group’s cfFrag scores were significantly higher than the HER2-positive group (Wilcoxon, *P* = 4.1×10^−3^; Figure 3F and Figure S13). This suggested the potential relation between the cfDNA fragmentomics features and the molecular subtypes.

#### The cfDNA fragmentomics (cfFrag) model demonstrates robust performance in the down-sampling process and technical replicates

To decrease the potential cost and required blood samples in the future, we assessed the cfFrag model’s performance using downsampled WGS data (5 – 1×) in the validation cohort with five technique repeats generated for each coverage depth. The fragmentomics model maintained its predictive power during the down-sampling process without showing a significant decrease in AUCs, even at a depth of 3× (*P* > 0.05; **Figure 4A**).

**Figure 4.**
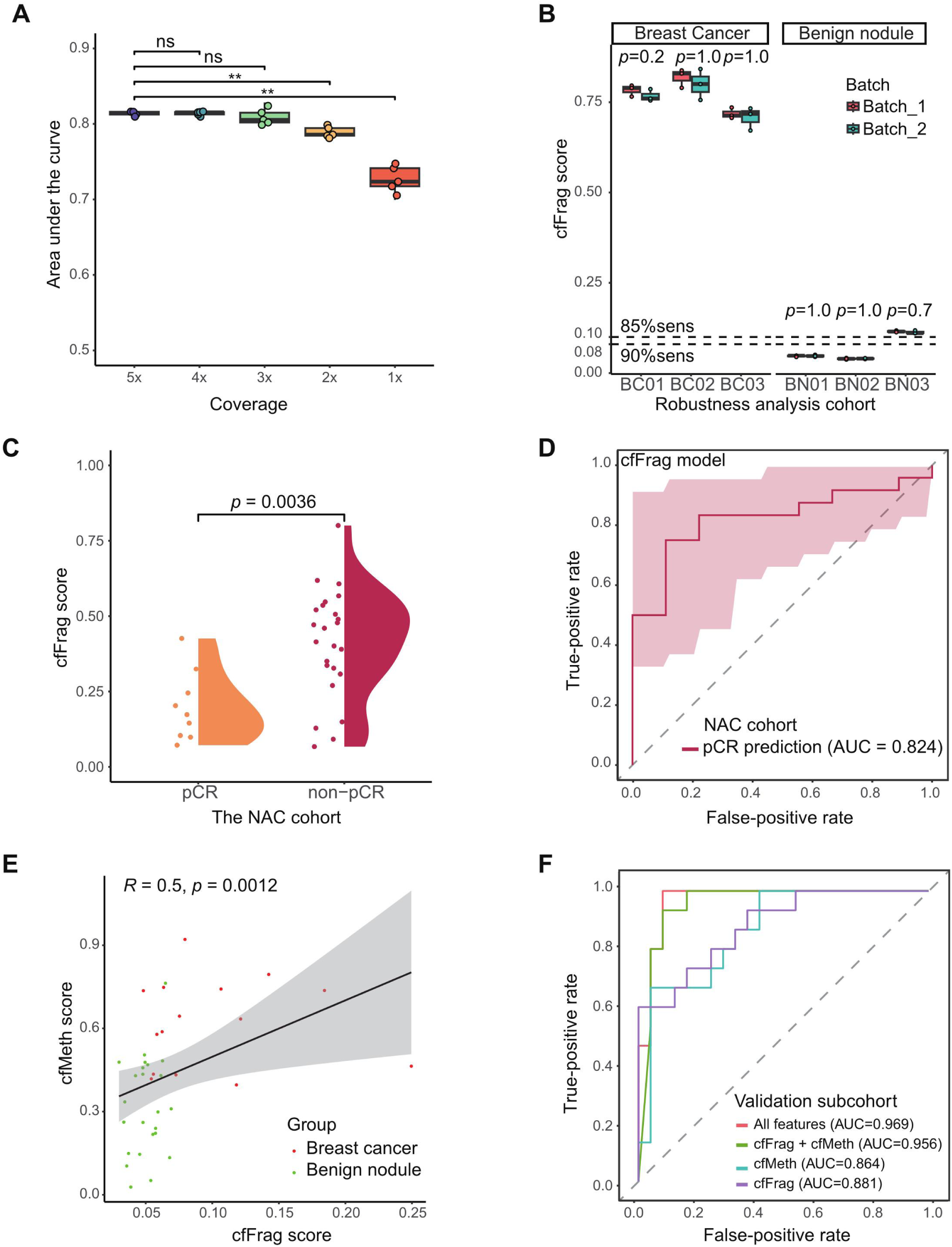
Comprehensively Evaluating cfFrag Model Using Additional Cohorts. **A.** Box plot for the validation cohorts’ AUCs for the down-sampling process (5× to 1×). There is no significant performance drop till 3× coverage (ns, no significance; ** *P* <0.01). **B.** Box plot for cfFrag scores in external test cohort containing 3 breast cancer patients and 3 benign nodule patients. For each patient, two batches × three repeats were performed. **C.** ROC curve for distinguishing patients with pathological complete response (N = 9) from patients without pathological complete response (N = 24) in a neoadjuvant therapy cohort. **D.** Violin plot using cfFrag cancer risk scores in patients with pCR and non-pCR. **E.** Scatter plot showing the correlation between cfMeth scores and cfFrag scores for a subset of patients in the validation cohort (N = 39) with previously reported whole-genome bisulfite sequencing data [4]. **F.** ROC curves for a subset of patients in the validation cohort (N = 39) with previously reported whole-genome bisulfite sequencing data [4] and imaging data using leave-one-out cross-validation. BC, breast cancer; BN, benign nodule; sens, sensitivity; NAC, neoadjuvant chemotherapy; pCR, pathological complete response; AUC, area under the curve.

To assess the robustness of the cfFrag model within and between runs, two batches of 10ml peripheral blood samples were collected from the six patients with a median interval of five days. The plasma samples were separated into three equal proportions as technical replicates for each batch, resulting in 36 samples. The model extracted and evaluated the fragmentomics profiles of these 36 samples. All three fragmentomics profiles (CNV, FSR, and FSD) showed no significant differences between the technical replicates and the two batches (Figure S14). Additionally, the robustness analysis showed no significant differences between runs and within runs (Wilcoxon, *P* = 0.2 – 1.0; Figure 4B).

#### The cfDNA fragmentomics (cfFrag) model can also predict the therapeutic pathological response for breast cancer patients after neoadjuvant chemoradiotherapy

To expand the clinical utility of the cfFrag model to predict the treatment response, we applied this assay to 33 female breast cancer patients receiving the NAC. The cfDNA fragmentomics profiles were generated using post-NAC plasma samples and were subsequently predicted by the cfFrag model. As a result, the cfFrag scores for pCR patients were significantly lower than the non-pCR patients (Wilcoxon, *P* = 3.6×10^−3^; Figure 4C). However, it is noted that the cfFrag scores for the pCR patients were still higher than those for patients with benign nodules. Moreover, the cfFrag model demonstrated excellent performance in distinguishing between patients with pCR and with non-pCR, yielding an AUC of 0.82 (95% CI: 0.68 – 0.97; Figure 4D). This indicated that the cfFrag model has the potential to predict the therapeutic response and minimal residual disease for post-NAC breast cancer patients.

#### The fragmentomics and methylation of cfDNA exhibit complementarity in breast cancer detection

To investigate the potential use of combined WGS with whole-genome bisulfite sequencing (WGBS) data for the differentiating power of breast cancers and benign nodules. We selected 39 patients from the prospective validation cohort (including 15 breast cancers and 24 benign nodules) enrolled in a methylation-based breast cancer early detection analysis to generate the breast cancer risk score (the cfMeth score) through the WGBS [4]. We found that the cfFrag and cfMeth scores were positively correlated (Spearman’s rank correlation coefficient, *R* = 0.5, *P* = 1.2×10^−3^; Figure 4E). Due to the limited size, leave-one-out cross-validation was performed. The combined (cfFrag + cfMeth) model showed better performance (AUC=0.96, 95% CI: 0.89 – 1.00) than the cfFrag model alone (AUC=0.88, 95% CI: 0.77 – 0.99) and the cfMeth model alone (AUC=0.86, 95% CI: 0.75 – 0.98), while the addition of imaging data (cfFrag + cfMeth + Xray + Ultrasound) further improved the performance (AUC = 0.97, 95% CI: 0.92 – 1.00; Figure 4F).

#### The joint diagnostic model combining the cfDNA fragmentomics (cfFrag) scores and breast imaging shows superior performance in detecting breast cancer

To further improve the performance of the cfDNA fragmentomics-based approach cost-effectively, a joint diagnostic model was constructed by integrating the cfFrag scores and the breast imaging reporting and data system (BI-RADS) categories for mammography and ultrasound using the machine-learning process. As a result, the joint model risk score exhibited a significant difference between the breast cancer and benign nodule groups in both the training and validation cohorts (Wilcoxon, *P* < 2.2×10^−16^; **Figures 5A and 5B**).

**Figure 5.**
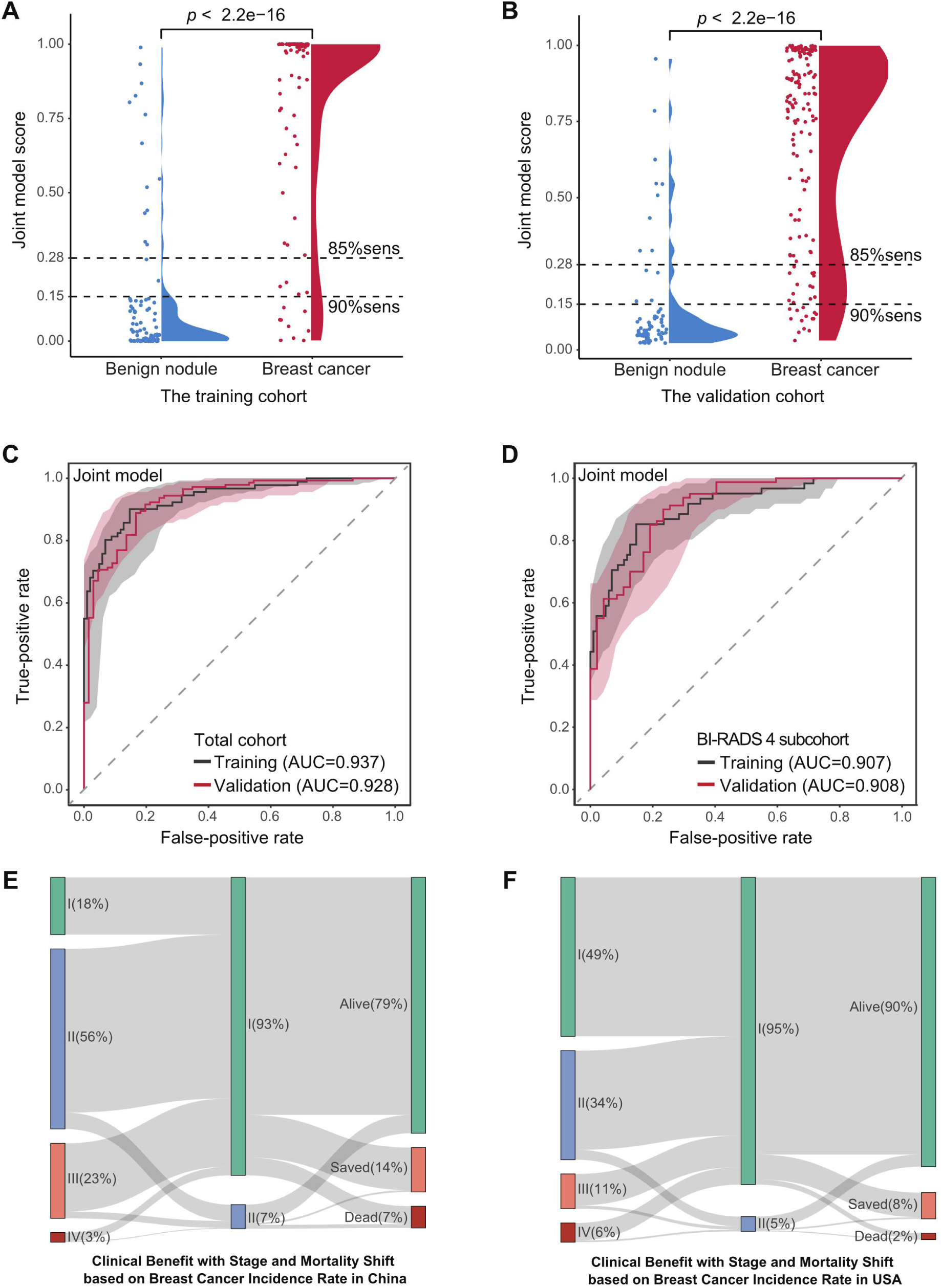
Evaluating Performance for Joint Model Using Both Fragmentomics and Imaging Techniques. **A.** Violin plots illustrating cancer score distribution of the joint model in the benign nodule and breast cancer groups in the training cohort. **B.** The score distribution of the joint model in the benign nodule and breast cancer groups in the independent validation cohort. **C.** ROC curves using the training cohort (5-fold cross-validation) and the independent validation cohort. **D.** ROC curves for subset patients with BI-RADS category 4 in the training cohort (5-fold cross-validation) and the validation cohort. **E.** Potential clinical benefit evaluation using breast cancer statistics in China. **F.** Potential clinical benefit of the joint model in the USA. The left bars show the current stage distributions of newly diagnosed breast cancer, and the middle bars indicate the stage distributions for potential clinical utilization of the joint model in the two countries. Accordingly, mortality shifts and 5-year survival benefits (orange bars) achieved by using the joint model are shown in the right bars. Abbreviation: sens, sensitivity; BI-RADS, breast imaging reporting and data system.

The joint diagnostic model showed superior performance for distinguishing breast cancers from benign nodules, with AUCs of 0.94 (95% CI: 0.90 – 0.97) and 0.93 (95% CI: 0.89 – 0.97) in the training and validation cohorts, respectively (Figure 5C), which was significantly higher than the cfFrag model alone, as well as the traditional mammography and ultrasound (all *P* < 0.05; Figure S14). Furthermore, the joint model could reach a high specificity of 80.3% (95% CI: 68.7 – 89.1%) at the designed 90.2% sensitivity (95% CI: 84.1 – 94.5%) in the independent validation cohort (**Table 2**). Furthermore, the joint model maintained its performance within women with the BI-RADS 4 lesions, reaching AUCs of 0.91 (95% CI: 0.86 – 0.95) and 0.91 (95% CI: 0.86 – 0.96) in the training and validation cohorts, respectively (Figure 5D).

**Table 2.** Evaluating the Joint Model Performances in the Training and Validation Cohorts.

|  | Training cohort<br>(5-fold cross-validation) | Prospective validation cohort |
| --- | --- | --- |
| Breast cancer patients (n) | 91 | 143 |
| Benign nodule patients (n) | 102 | 66 |
| Sensitivity (95% CI) | 90.1% (82.1-95.4%) | 90.2% (84.1-94.5%) |
| Specificity (95% CI) | 85.3% (76.9-91.5%) | 80.3% (68.7-89.1%) |
| PPV (95% CI) | 84.5% (75.8-91.1%) | 90.8% (84.9-95.0%) |
| NPV (95% CI) | 90.6% (82.9-95.6%) | 79.1% (67.4-88.1%) |
| Accuracy (95% CI) | 87.6% (82.1-91.9%) | 87.1% (81.8-91.3%) |

Abbreviation: CI, confidence interval; PPV, positive predictive value; NPV, negative predictive value.

#### The joint model illustrates the potential for increased early detection rates and improved survival outcomes in China and the USA

To assess the potential clinical benefits of the joint model in a real-world setting, we utilized an intercept model developed by Hubbell *et al* [24]. Currently, only 18% of breast cancer patients are diagnosed at stage I in China. By utilization of the joint model, the detection rate of stage-I breast cancer could be elevated to 93%. Accordingly, less breast cancer would be diagnosed at stages II-IV. Based on the stage shifts, it was estimated that the joint model could increase the 5-year survival rates of breast cancer in China by 14% (Figure 5E). Similarly, in the USA, the increased detection rate of stage-I breast cancer (95%) and the 5-year survival benefit (8%) are also estimated (Figure 5F).

## Discussion

Current imaging-based breast cancer screening methods suffer from high false positives and inconclusive results among female patients with benign breast nodules, which leads to intrusive biopsy and unnecessarily adds to the discomfort. In our study, we provided a highly sensitive, non-invasive diagnostic tool for early-stage breast cancer detection through the blood-based cfDNA fragmentomics analysis, especially against control patients with radiographically malignant-suspicious yet pathologically benign breast nodules. Notably, we have demonstrated a significant complementarity between cfDNA fragmentomics, traditional imaging, and cfDNA methylation features in the early detection of breast cancer and the assessment of the benign or malignant nature of breast nodules. The combination of cfDNA fragmentomics and traditional imaging findings, as well as the combination of cfDNA fragmentomics and cfDNA methylation features, can further enhance the diagnostic accuracy of breast cancer, aligning with our previous findings on combining cfDNA methylation and traditional imaging in breast cancer. The joint diagnostic model, integrating our non-invasive cfDNA fragmentomics assay with image findings, achieves high diagnostic accuracy (AUCs=0.93 – 0.94). Accordingly, the joint model can guide biopsy decisions and reduce unnecessary invasive interventions by 80.3-85.3% in patients with suspicious imaging results.

The detection rate/sensitivity is crucial to avoid cancer diagnostic delay. Thus, 85-90% sensitivities for early-stage breast cancer were set as the primary endpoint for the fragmentomics and joint models in this study. The fragmentomics-only and joint models performed robust detection rates in early-stage breast cancer, even in women with small nodules or inconclusive imaging results (BI-RADS 4 lesions). Due to the potential stage shift and increase in the stage-I breast cancer diagnosis rate, our joint model was estimated to save a significant number of breast cancer patients (an extra 8 – 14%) in the USA and China. However, further real-world studies are still needed to identify the cut-off value for each model with the best cost-effectiveness in different populations.

The detection rates were robustly elevated across increasing breast cancer stages in this study. Intriguingly, the cfDNA fragmentomics signal was more significant in patients with DCIS than those with stage-I breast cancer, which is consistent with our methylation-based cfDNA analysis [4] but inconsistent with previous mutation-based cfDNA analysis [25]. It indicates the advantages of epigenetic-based and fragmentomics-based cfDNA analysis in the early detection of DCIS/stage-0 breast cancer. In addition, the cfFrag model has shown sufficient specificity in asymptomatic healthy women, further indicating the potential clinical utility of the cfFrag model in population-based breast cancer screening.

The use of peripheral cfDNA has gained prominence in early cancer detection, as methylation-based and fragment-based cfDNA markers have demonstrated effectiveness in detecting many cancer types [4, 13–15, 26]. Methylation features in cfDNA are related to the cancer tissue-of-origin, while cfDNA fragmentomics features are linked to the abnormal DNA nuclease activities in cancers [27]. Compared to the methylation-based cfDNA approach, fragment-based cfDNA assays offer advantages such as lower cost by avoiding sodium bisulfite treatment and requiring less blood sample volume because of low sequence depth. Interestingly, our combined analysis of cfDNA methylation and fragmentomics reveals that combining the fragmentomics (cfFrag score) and the methylation markers (cfMeth score) could achieve superior performance than each marker separately, which was in agreement with the result of the recent sub-study in the Circulating Cell-free Genome Atlas [28].

Monitoring treatment response is crucial to deciding the subsequent treatment strategies for breast cancer patients receiving NAC, but this was unmet using the current methods [29]. Recently, mutation-based and methylation-based ctDNA detection approaches have been demonstrated to predict the treatment response and residual disease in post-NAC breast cancer patients [30, 31]. Our study suggests that the features of cfDNA fragmentomics could be used as an alternative approach to evaluate the treatment response in breast cancer patients.

### Limitations

Firstly, the sample size of the combining analysis of the cfDNA fragmentomics and methylation is relatively small. Multi-omics cfDNA analysis with large sample sizes is still needed to identify the optimal non-invasive combination with low cost using a trace amount of blood sample for the early detection of breast cancer. Secondly, similar to most previous cfDNA fragmentomics studies that only focused on one cancer type [13–15], we also aimed to identify the breast cancer-specific cfDNA fragmentomics features in this study. With the identification of the cfDNA fragmentomics spectrum in different cancer types in the future, it would be cost-effective to develop a pan-cancer diagnostic model to detect multiple types of cancer and indicate the cancer origin.

## Conclusions

This pilot study systematically evaluates performance in applying cfDNA fragmentomics as a non-invasive biomarker for breast cancer. The low-depth cfDNA fragmentomics profiling with automated machine learning demonstrated excellent and robust performance in distinguishing early-stage breast cancers from benign nodules with inconclusive imaging results, the predictive value of NAC response, and sufficient specificity in asymptomatic healthy women in a multi-center prospective setting. The combination of non-invasive cfDNA fragmentomics features and standard diagnostic imaging improved the rate of accurate detection of early breast cancer. This approach holds promise for improving clinical outcomes and streamlining healthcare practices.

## Materials and methods

### Study Design and Participants

In this multi-center study, we recruited female patients independently in three centers in China. The training set enrolled 200 consecutive female patients with malignant-suspicious breast imaging results from the YYH in Yantai. The external independent validation sets, referred to as Beijing cohorts, prospectively enrolled 209 consecutive female patients who underwent breast lesion biopsy, 33 female breast cancer patients after NAC, and six female patients with repeating samples for robustness analysis from the CHCAMS in Beijing. The external screening cohort recruited 119 asymptomatic healthy women from our previous Nanjing cohort [14]. The recruitment period was from January 1, 2019, to August 1, 2022. This study adhered to the guidelines of the STARD (Standards for Reporting of Diagnostic Accuracy Studies).

### Sample Collection and Clinical Evaluation

We collected 10 ml of peripheral blood samples from each participant before biopsy or surgery. In the Yantai cohort (training set), the collected blood samples were kept in EDTA blood collection tubes (Becton Dickinson, CA) at a temperature of 4°C and underwent centrifugations (1,800 g for 10 minutes and 16,000 g for 10 minutes both at 4°C) within 2 hours. In the Beijing cohorts (independent validation set), the collected blood samples were kept in the CELL-FREE DNA BCT^®^ blood collection tubes (Streck, NE) at room temperature (RT, 15 – 25°C). Plasma was extracted within 48 hours following blood collection by centrifugations of the blood according to the protocols in the training set.

Standard mammography and ultrasonography techniques were conducted at two centers and independently interpreted according to the BI-RADS standard. Patients with suspicious breast imaging results underwent surgical or core needle biopsies. The pathological examination of tissue specimens confirmed the malignant or benign status of each participant. Women with negative imaging or biopsy results were excluded from having breast cancer after a 6-month follow-up. The molecular subtype of each lesion was determined according to the pathologic criteria for HR (including estrogen receptor and progesterone receptor) and human epidermal growth factor receptor 2 (HER2) [32].

### Library preparation and whole genome sequencing

For the cfDNA extraction, we used the liquid handling platform (Hamilton Microlab STAR, Hamilton Company, NV) and the QIAamp Circulating Nucleic Acid Kit (Qiagen, Germany) according to previously reported protocols [13]. The Qubit dsDNA HS Assay Kit (Thermo Fisher Scientific, MA) was then utilized for measuring the extracted cfDNA’s concentration. The PCR-free WGS library was automatically constructed on Biomek (Beckman Coulter, UK), using 5-10 ng of cfDNA sample and the KAPA Hyper Prep Kit (KAPA Biosystems, MA). The constructed library was quantified by the KAPA SYBR FAST qPCR Master Mix (KAPA Biosystems, MA) before paired-end sequencing on NovaSeq platforms (Illumina, CA).

For the quality control of bioinformatics analysis, Trimmomatic [33] was used to trim the raw sequencing data. The removal of PCR duplicates was performed by the Picard toolkit (http://broadinstitute.github.io/picard/). The high-quality reads were then mapped to the human reference genome (GRCh37/UCSC hg19) using BWA sequence aligner [34].

### Fragmentomics profiling

As tumor cell fragments are shorter than those from normal cells [35], the FSR profile analyzes the ratio of short fragments in the human genome. Short fragments are defined as 100 – 150bp and long fragments are defined as 151 – 220bp [13, 15, 20]. The human genomes were divided into 5Mb bins, in which the ratios of short to long fragments were calculated, resulting in a total of 1,082 (541 bins × 2) FSR features. The FSD profile focused on the detailed length patterns of cfDNA fragments, categorizing these fragments based on increments of 5bp in the range of 100bp to 220bp [13, 36]. The proportion of fragments in each bin was computed on the human chromosome arm level for human autosomes, resulting in 936 FSD features that machine learning algorithms can employ. The CNV profile was calculated using ichorCNA [15]. For each sample, the genome was divided into 1Mb bins. The depth for each bin was then compared to the default baseline using the Hidden Markov Model (HMM). The log2 ratio for each bin was calculated, generating 2475 features. The profiling of Griffin, neomer, and MBP was present in **Supplementary methods** (File S1).

### cfFrag Model construction and validation

A machine-learning process that utilizes five different algorithms, including the random forest, the generalized linear model, the deep learning, the gradient boosting machine, and the eXtreme gradient boosting [13], was employed to generate optimal base learners. A breast cancer prediction model, namely the cfFrag scores, was developed using the mean value of top base learners ranked by the AUC of the 5-fold cross-validation for the optimal three fragmentomics profiles in the training cohort (Yantai cohort; File S1).

The machine-learning process was also utilized to develop a joint diagnostic model for the training cohort, which adopts the cfFrag scores and the BI-RADS categories of mammography and ultrasound. A similar automated machine-learning process was used to construct the joint diagnostic model by using the cfFrag scores as numeric features and the BI-RADS by mammography and ultrasound as categorical features. The process utilized a randomized search for automatic algorithm selection and hyperparameter tuning. The best-performing model was selected from a total of 200 trained models based on the highest AUC using the training cohort via a 5-fold cross-validation approach. Cut-off values were determined using a 5-fold cross-validation to predict the score of the training cohort to reach 85% and 90% sensitivities. The external independent validation cohorts evaluated the joint diagnostic models’ performance. In addition, to expand the clinical utility of the cfFrag model, its performance was further evaluated in the NAC cohort.

### Statistical Analysis

The receiver operating characteristic (ROC) curves were generated by the pROC package (version 1.17.0.1) [37]. The sensitivity, specificity, and accuracy with the corresponding 95% confidence intervals (CI) were calculated by the epiR package (version 2.0.19) [38]. Propensity score matching analysis used the MatchIt package (version 4.2.0) [39]. All statistical analyses, including student’s *t*-test, Wilcoxon, and ANOVA, were performed in R (version 3.6.3) [40].

## Ethical statement

This study was reviewed and approved by the ethics committees of each center (22/291-3493 for the CHACMS and 2020-289 for the YYH). Each participant provided written informed consent.

## Data and code availability

Raw data from the deidentified participant and analytic code generated in this study are available for non-commercial use upon reasonable request to Dr. Jiaqi Liu.

## Acknowledgments

This research was funded in part by the National Natural Science Foundation of China (82272938 to Jiaqi Liu), the Beijing Nova Program (20220484059 to Jiaqi Liu), and the CAMS Innovation Fund for Medical Sciences (2021-I2M-1-014 to Jiaqi Liu). This study is part of the DETEct study (Deciphering Epigenetic signatures in Tumor and Exploiting ctDNA). We thank all the individuals involved in the study for their participation.

## Supplemental material

**File S1. Supplementary methods**

**Figure S1. Multi-omic Cell-free DNA Profiles of Breast Cancer Patients, and Subjects with Benign Nodule.**

**A.** Frequencies of chromosome arm-level copy number variations (CNVs) in subjects with breast cancers and benign nodules. Amplifications are represented in red, while losses are depicted in blue. **B.** Fragment size distribution (FSD) in chromosome 18p across various groups. The distribution illustrates the fragment size profiles among subjects with benign nodules and breast cancers. **C.** Ratio of short (100-150bp) fragments to long (150-220bp) fragments across all 5Mb bins on chromosomal arms in subjects with breast cancers and benign nodules. Abbreviation: CNV, copy number variation; BC, breast cancer; BN, benign nodule.

**Figure S2. The ichorCNA Tumor Fraction Distribution of Breast Cancer and Benign Nodule in the Training Cohort and the Validation Cohort.**

We used the ichorCNA reported tumor fraction (TF) to show the differences in CNV profile between breast cancer (BC) patients and benign nodule (BN) patients. The TF by ichorCNA was significantly higher for the BC patients compared to the BN patients in both the training cohort (*P* = 8×10^−5^) and the validation cohort (*P* = 0.029). This suggests that while the BC and BN groups both vary substantially from health baselines, there are still distinguishable differences between the two groups. Abbreviation: TF, tumor fraction; CNV, copy number variation; BC, breast cancer; BN, benign nodule.

**Figure S3. The Area Under the Curve Distribution of Base Learners in the Training Cohort.**

A total of 24 (3×8) top base learners were selected to create the final cfFrag score by the 5-fold cross-validation AUC in the training cohort. Abbreviation: AUC, area under the curve.

**Figure S4. ROC Curves for Selected Base Learners in the Training Cohort.**

Among the three feature types, CNV showed the highest mean AUC of 0.742 [0.661 – 0.791] for its top 8 base learners, while the FSD and FSR showed similar predict power in mean AUC (0.706 [0.631-0.750] and 0.706 [0.647-0.754]), as shown in Table S3. Abbreviation: CNV, copy number variation; AUC, area under the curve; FSD, fragment size distribution; FSR, fragment size ratio.

**Figure S5. Feature Recursive Feature Elimination Analysis in the Training Cohort and the Validation Cohort.**

The cfFrag model showed possible overfitting in the training cohort using only subsets of top-performing features in the final model. The 5-fold cross-validation AUCs in the training cohort showed a gradual decrease as more features were used in the model construction process. Abbreviation: AUC, area under the curve.

**Figure S6. The cfFrag Score Distribution in the Training Cohort (5-fold cross-validation).**

**Figure S7. ROC Curve of the cfFrag Model in Subsets of Age-matched Patients and Patients with Small Nodule (≤1cm) in the Training and Validation Cohorts.**

**A.** Receiver operating characteristic (ROC) curves of the cfFrag model using an age-matched subset in the training cohort (5-fold cross-validation) and the independent validation cohort. **B.** ROC curves of the cfFrag model using an age-matched subset in the training cohort (5-fold cross-validation) and the independent validation cohort. The shadow areas indicate the 95% confidence intervals (CI). Abbreviation: ROC, receiver operating characteristic; AUC, area under the curve; CI, confidence interval.

**Figure S8. ROC Curves for Traditional Imaging Technique in Different Cohorts.**

Abbreviation: BI-RADS, Breast Imaging Reporting and Data System; ROC, receiver operating characteristic; AUC, area under the curve.

**Figure S9. Performance Evaluation for Fragmentomics Model in Different Subgroups of Breast Cancer Patients in the Validation Cohort.**

Abbreviation: HR, hormone receptor; HER2, human epidermal growth factor receptor 2; TNBC, triple-negative breast cancer; BI-RADS, Breast Imaging Reporting and Data System.

**Figure S10. Performance Evaluation for Fragmentomics Model in Different Subgroups of Patients with Benign Nodule in the Validation Cohort.**

Abbreviation: BI-RADS, Breast Imaging Reporting and Data System.

**Figure S11. Bootstrapped (100 times) Performance Evaluation of Fragmentomics Model in Breast Cancer Subgroups in the Validation Cohort.**

The sensitivities derived from 100 bootstrap iterations for various breast cancer subgroups displayed patterns similar to our observations in Figure S5. Abbreviation: HR, hormone receptor; HER2, human epidermal growth factor receptor 2; TNBC, triple-negative breast cancer.

**Figure S12. Bootstrapped (100 times) Performance Evaluation of Fragmentomics Model in Benign Nodule Subgroups in the Validation Cohort.**

The specificities for the BN subgroup, assessed through 100 bootstrap iterations, align with the trends seen in the validation cohort (Figure S6).

**Figure S13. The cfFrag Score Distribution of Different Subgroups in the Validation Cohort (HR, HER2, and TNBC).**

Abbreviation: NS, no significance; HR, hormone receptor; HER2, human epidermal growth factor receptor 2; TNBC, triple-negative breast cancer.

**Figure S14. The Feature Correlation of Inter- and Intra-run Samples.**

No significant differences were observed between the technical replicates and the two batches in all three fragmentomics profiles, including copy number variation (A), fragment size distribution (B), and fragment size ratio (C). Abbreviation: CNV, copy number variation; FSD, fragment size distribution; FSR, fragment size ratio.

**Figure S15. Comparing the Joint Model against the Fragmentomics Model, Mammography, and Ultrasound.**

ROC curves for the training cohort (A; 5-fold cross-validation) and the prospective validation cohort (B). The Wilcoxon *P* values compare the joint model against each individual technique. Abbreviation: AUC, area under the curve.

**Table S1. Patient Characteristics.**

**Table S2. Selected top-performing base learners for constructing the final cfFrag model.**

**Table S3. CNV features ranked by their importance.**

**Table S4. FSR features ranked by their importance.**

**Table S5. FSD features ranked by their importance.**

**Table S6. Evaluating the Fragmentomics Model Performances at 85% Sensitivity Cutoff.**

