## Supplementary material for "Cell-free DNA Fragmentomics Assay to Discriminate the Malignancy of Breast Nodules and Evaluate Treatment Response": File S1 Supplementary methods

***Additional fragmentomics feature profiling***

To generate the Griffin feature, a profiling framework was adopted from Doebley and colleagues [1] to evaluate nucleosome occupancy and the corresponding protection of cell-free DNA (cfDNA). The GC-corrected coverage profile was quantified by employing three observable characteristics: the central coverage value measured at 30 base pairs from the specified location, the average coverage calculated within a 1000 base pair radius of the location, and the amplitude ascertained through the application of a Fast-Fourier Transform analysis. A total of 854 Griffin features representing transcription factor binding sites were generated from the low-pass whole-genome sequencing (WGS) data.

The neomer features were defined as short DNA sequences, which recur in tumor genomes but are absent from the human reference genome, by Georgakopoulos-Soares and colleagues [2]. A total of 977 recurrent single-nucleotide polymorphisms (SNPs) were identified from 2577 cancer patient samples using the PCAWG database (https://dcc.icgc.org/releases/PCAWG/). In total, 4,616 neomers of 16bp length were extracted from these SNPs, which were then filtered against common population variants compiled in the Genome Aggregation Database (gnomAD v2) [3], resulting in a final total of 1,758 neomer feature. The neomer features were profiled as the ratio of neomer-detecting reads over the total reads and the read count of each of the 1,758 neomers.

The motif breakpoint (MBP) feature [4] examined the frequencies of the 6bp motif at the 5’ breakpoints on the human reference genome hg19, which extended 3bp to each direction. A total of 4,096 ($4^6$) MBP features were generated from the low-pass WGS data.

***cfDNA fragmentomics (cfFrag) score construction***

An automated machine-learning (autoML) process that utilizes five different algorithms, including generalized linear model (GLM), gradient boosting machine (GBM), random forest (RF), deep learning (DL), and eXtreme gradient boosting (XGBoost) [5], was employed to generate optimal base learners. The autoML utilized a randomized search for automatic algorithm selection, as well as for hyperparameter tuning. For each cfDNA fragmentomics feature type, a total of 200 base learners were constructed using an autoML procedure, which performs hyperparameter tuning via random grid search.

The area under the curves was calculated using the training dataset via a 5-fold cross-validation approach for all base learners. For each feature type, the base learners were ranked by their AUCs, and the top 8 performing were then selected for constructing the cfFrag score. The final cfFrag score for each sample was then generated by calculating the mean predict score of the total 24 ( 3 × 8 ) optimal base learners.

$$cfFrag\ Score = \left(\frac{1}{8}\right) * \sum\left(Score(FSR_i)\right) + \left(\frac{1}{8}\right) * \sum\left(Score(FSD_i)\right) + \left(\frac{1}{8}\right) * \sum\left(Score(CNV_i)\right)$$

The base learner predicts the score, which ranges from 0 to 1, representing the probability of a sample being breast cancer (0 = perfect benign nodule, 1 = perfect breast cancer; CNV, copy number variation; FSD, fragment size distribution; FSR, fragment size ratio.).

***Fragmentomics features evaluation***

We further evaluated the contribution of individual fragmentomics features by ranking them according to their importance in the final cfFrag model. For each base learner, we calculated the relative importance of individual features and sorted them from highest to lowest importance (using the maximum rank method for any tied values). For each of the three feature types, including copy number variation (CNV), fragment size distribution (FSD), and fragment size ratio (FSR), we determined the final importance of individual features by ranking the summed importance ranks of the eight selected top-base learners. A lower summed importance rank indicates a higher feature importance in the final cfFrag model.
