## Supplementary figures and images for "Cell-free DNA Fragmentomics Assay to Discriminate the Malignancy of Breast Nodules and Evaluate Treatment Response"

### Figure S1

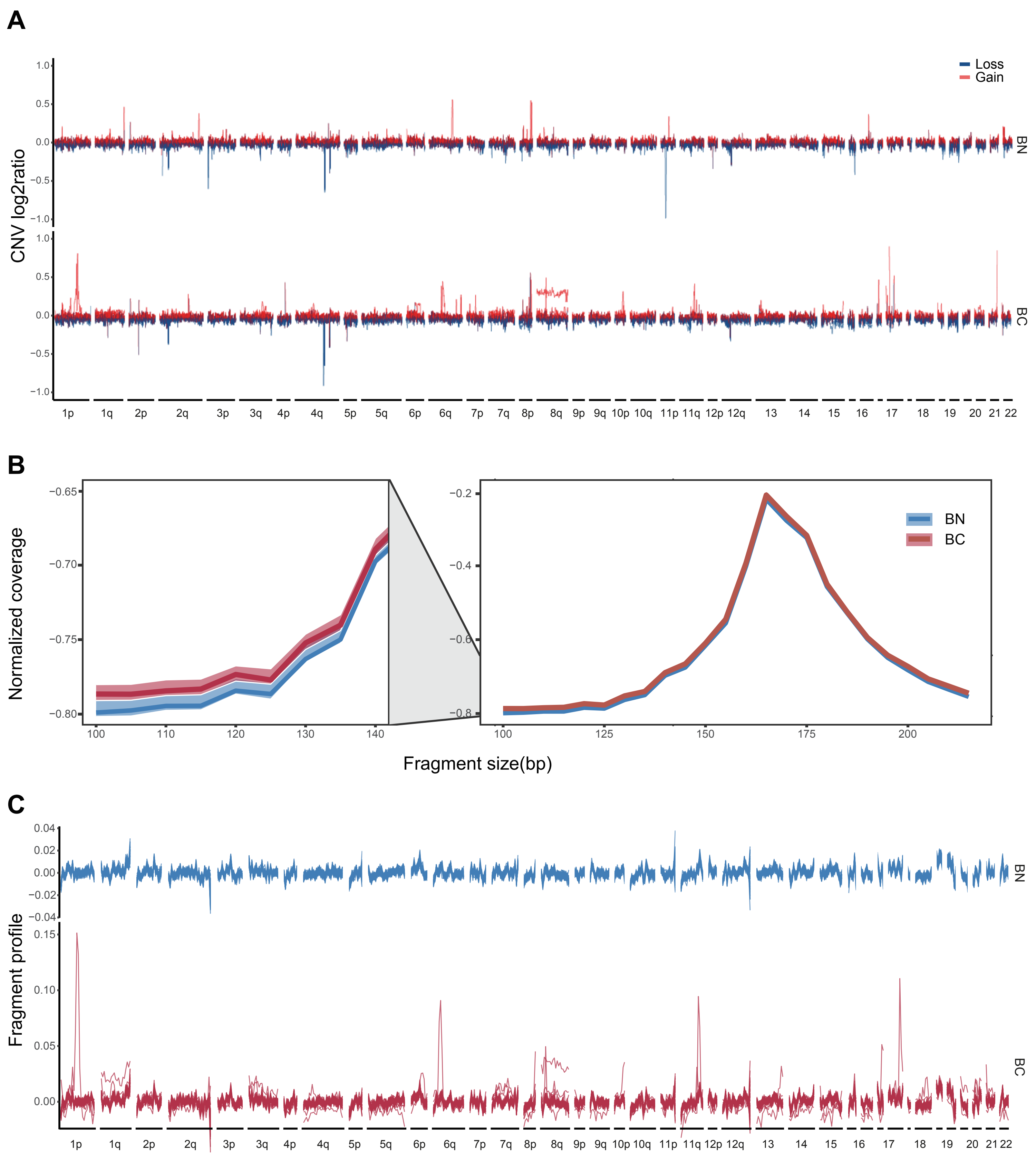

### Figure S2

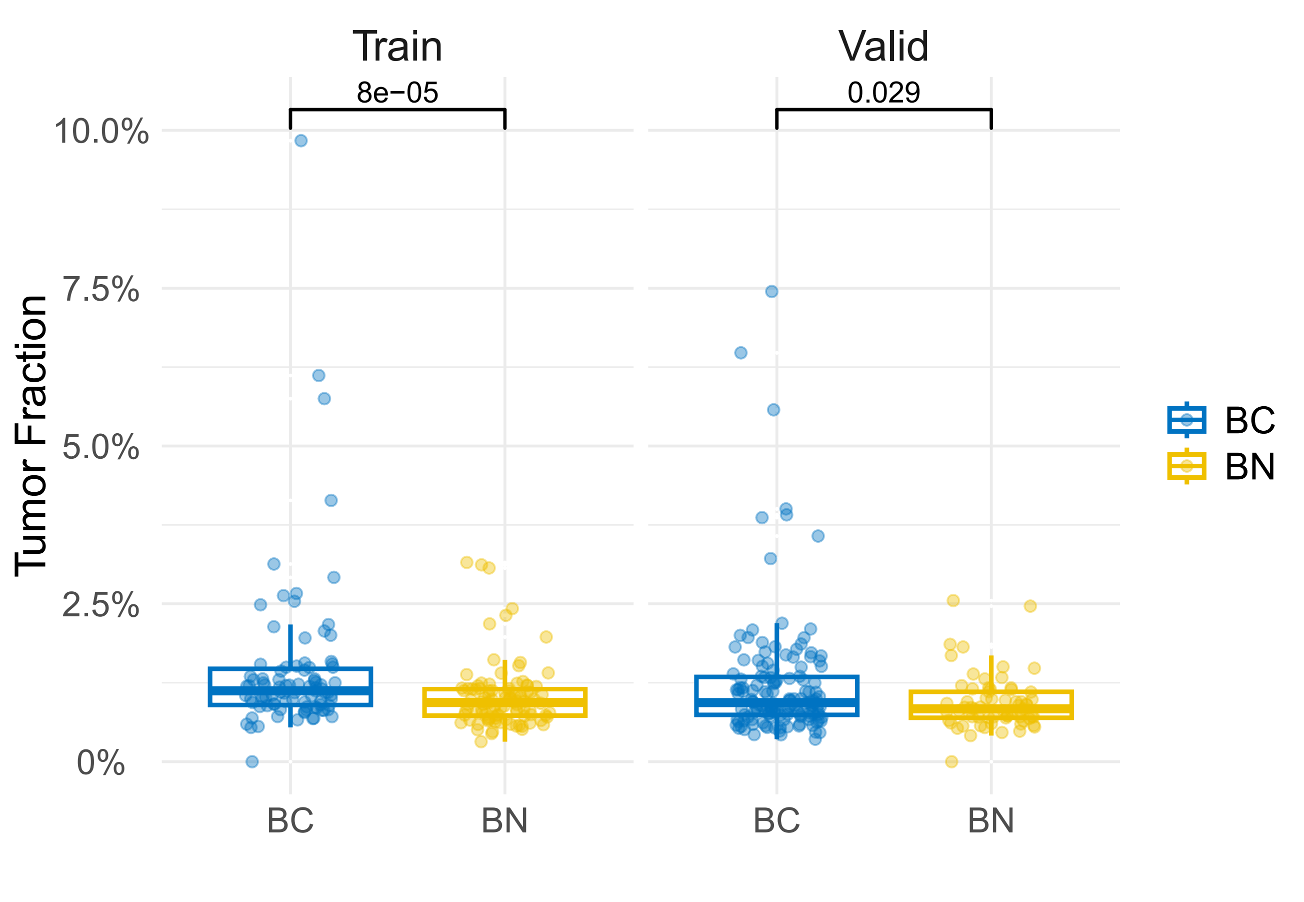

### Figure S3

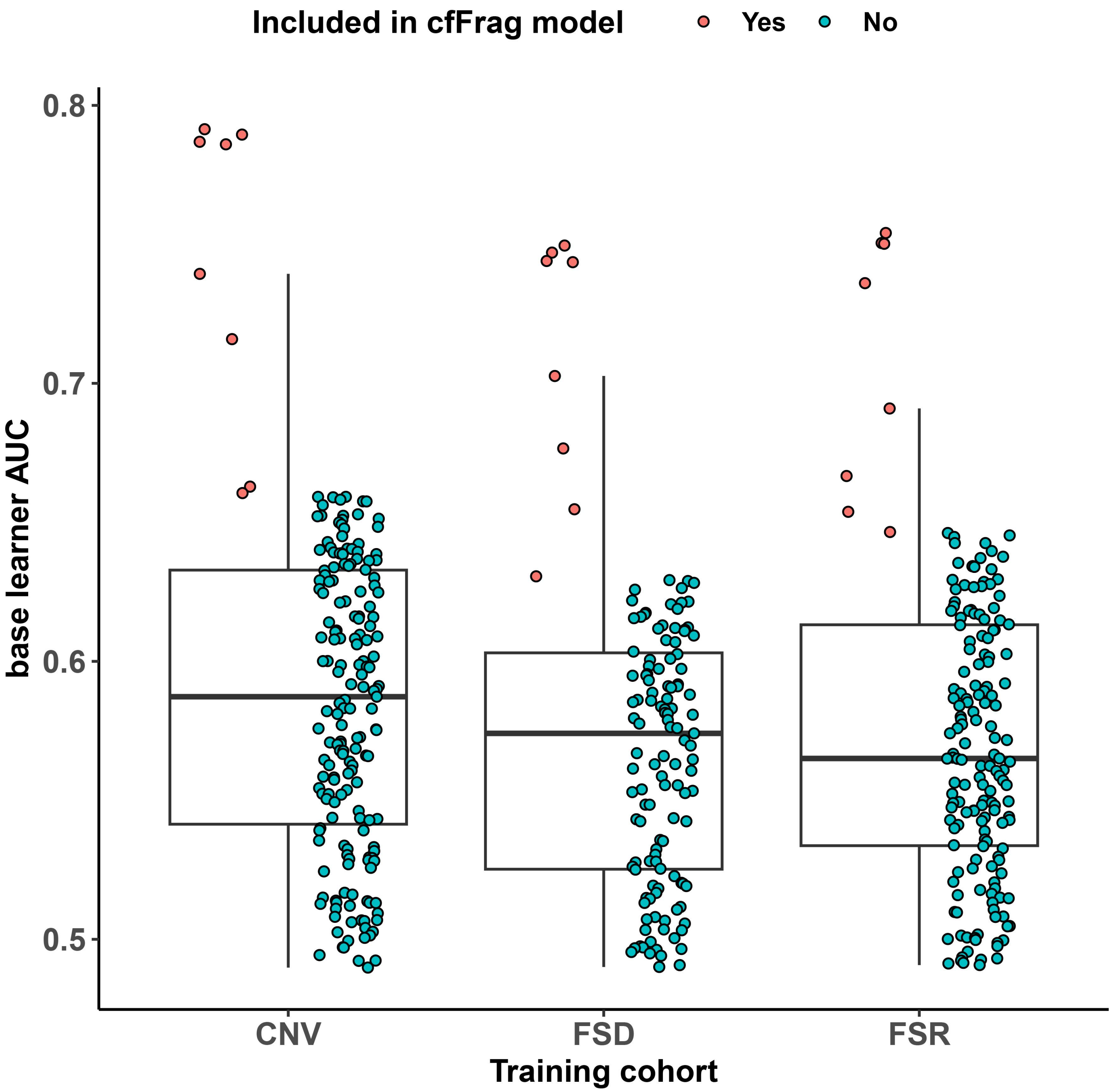

### Figure S4

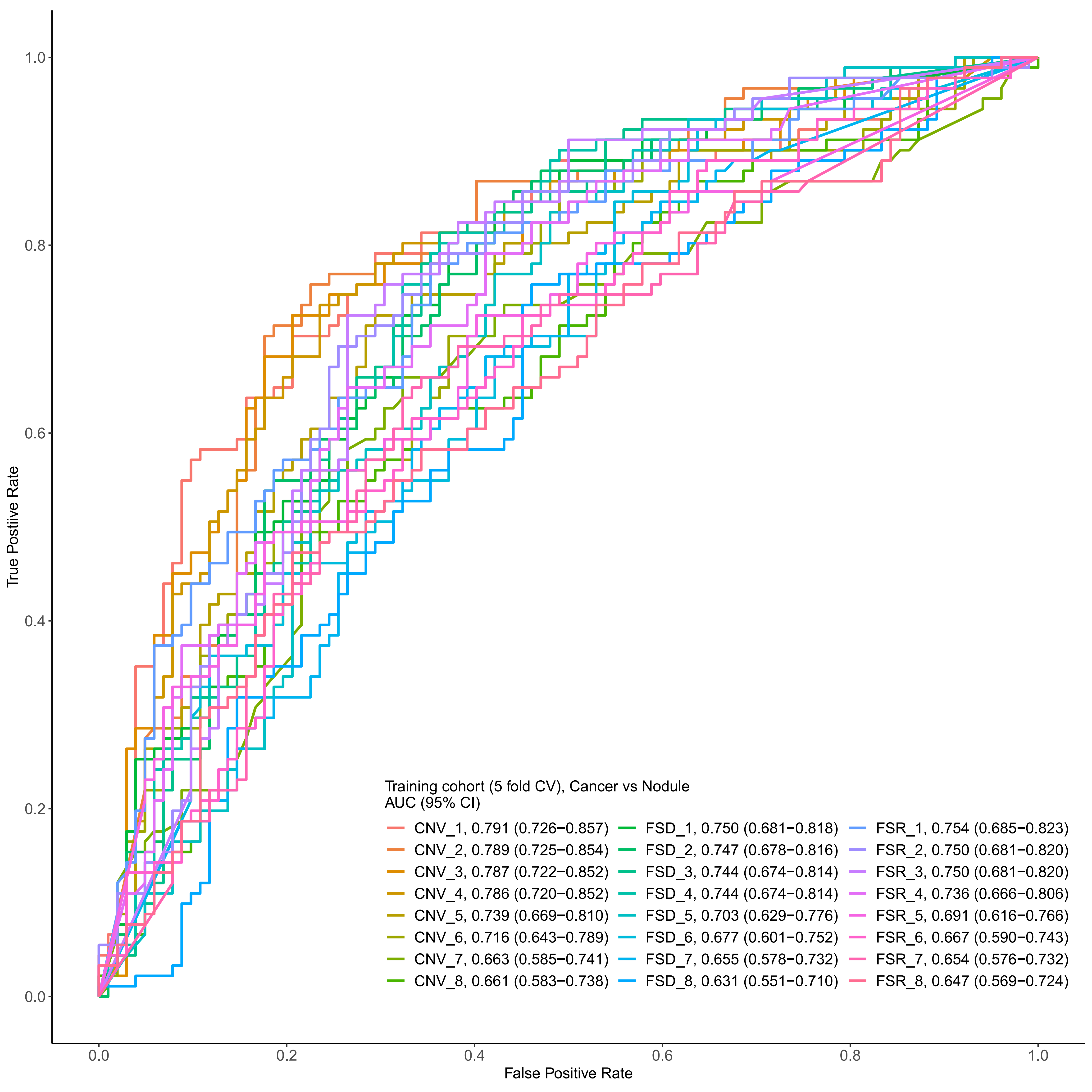

### Figure S5

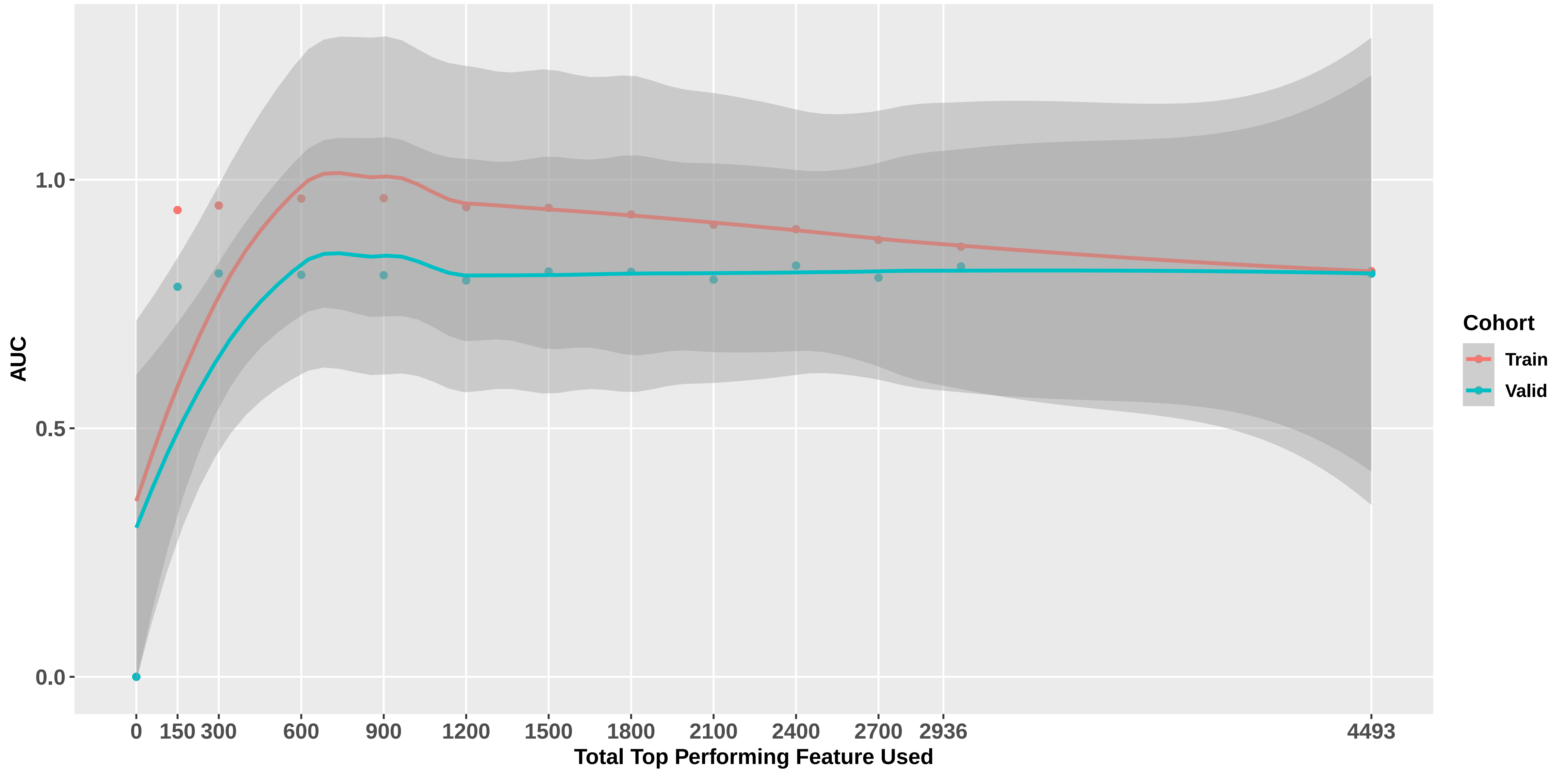

### Figure S6

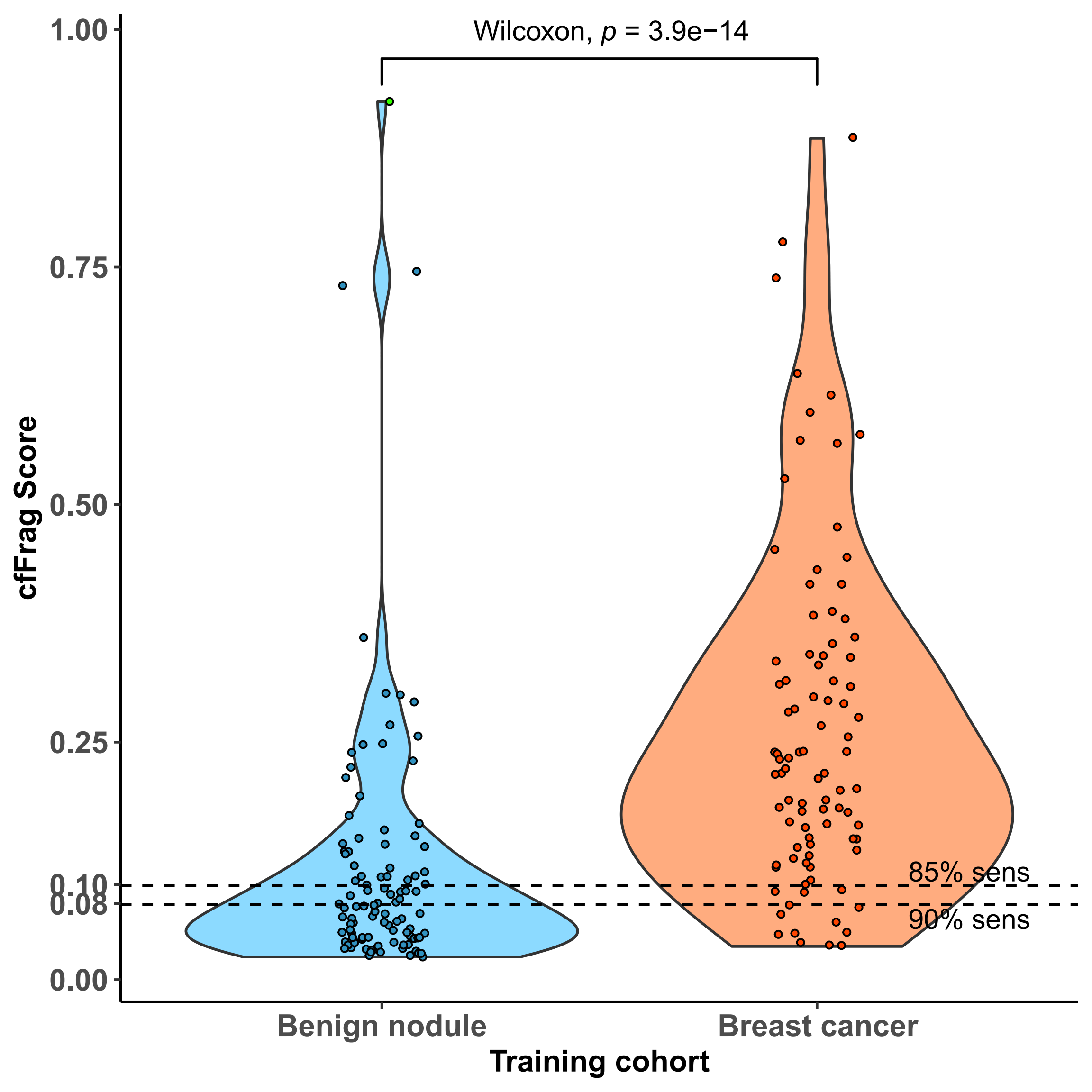

### Figure S7

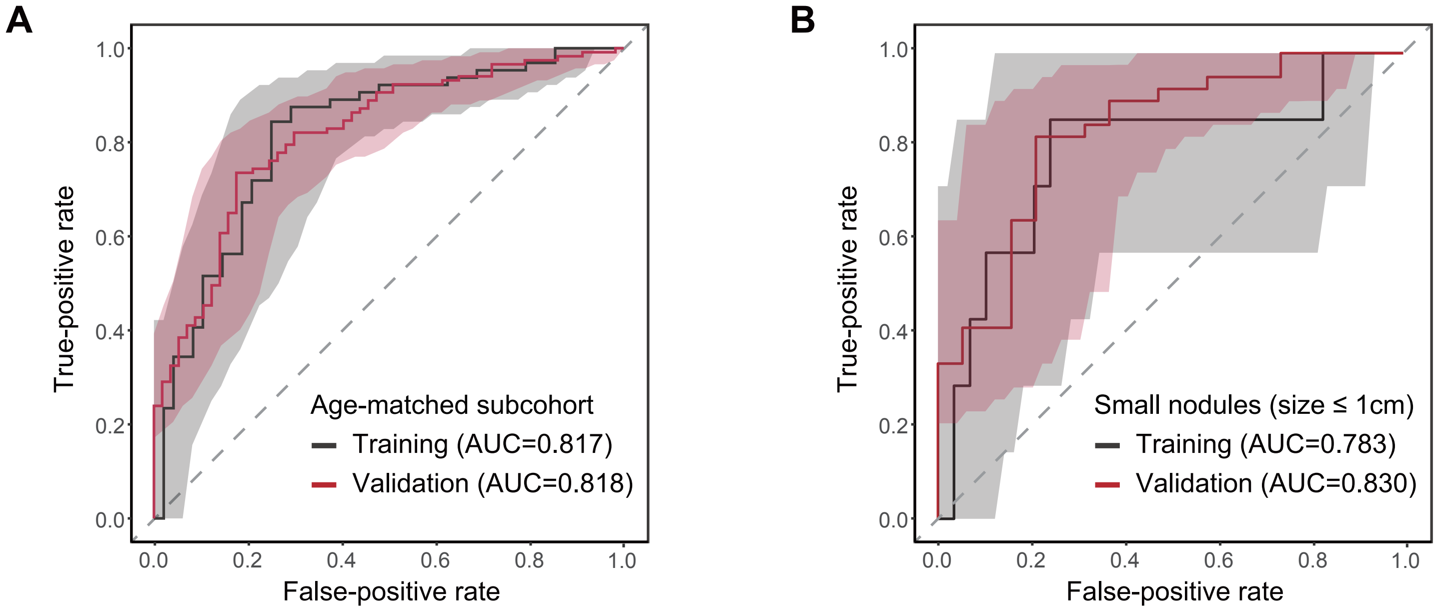

### Figure S8

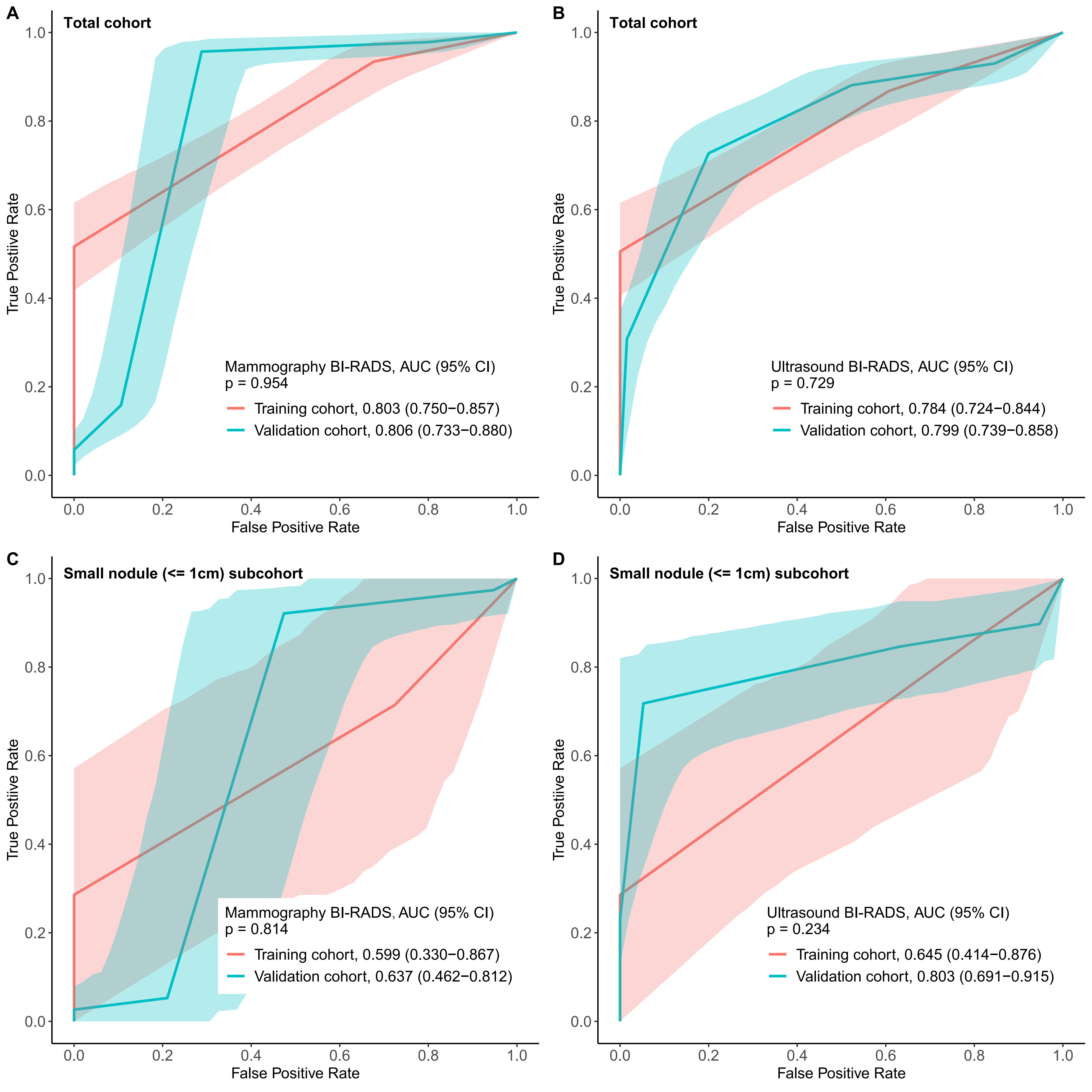

### Figure S9

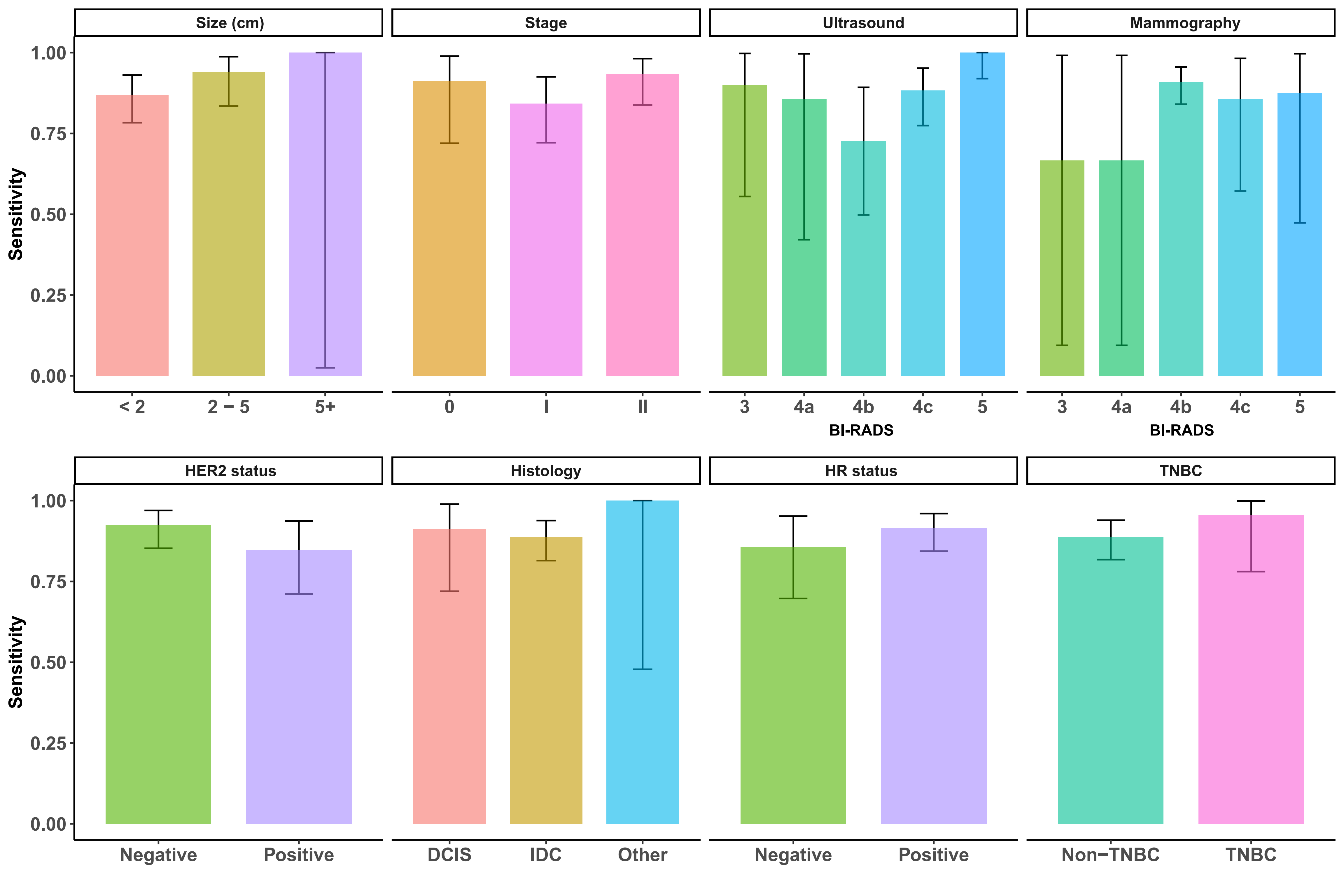

### Figure S10

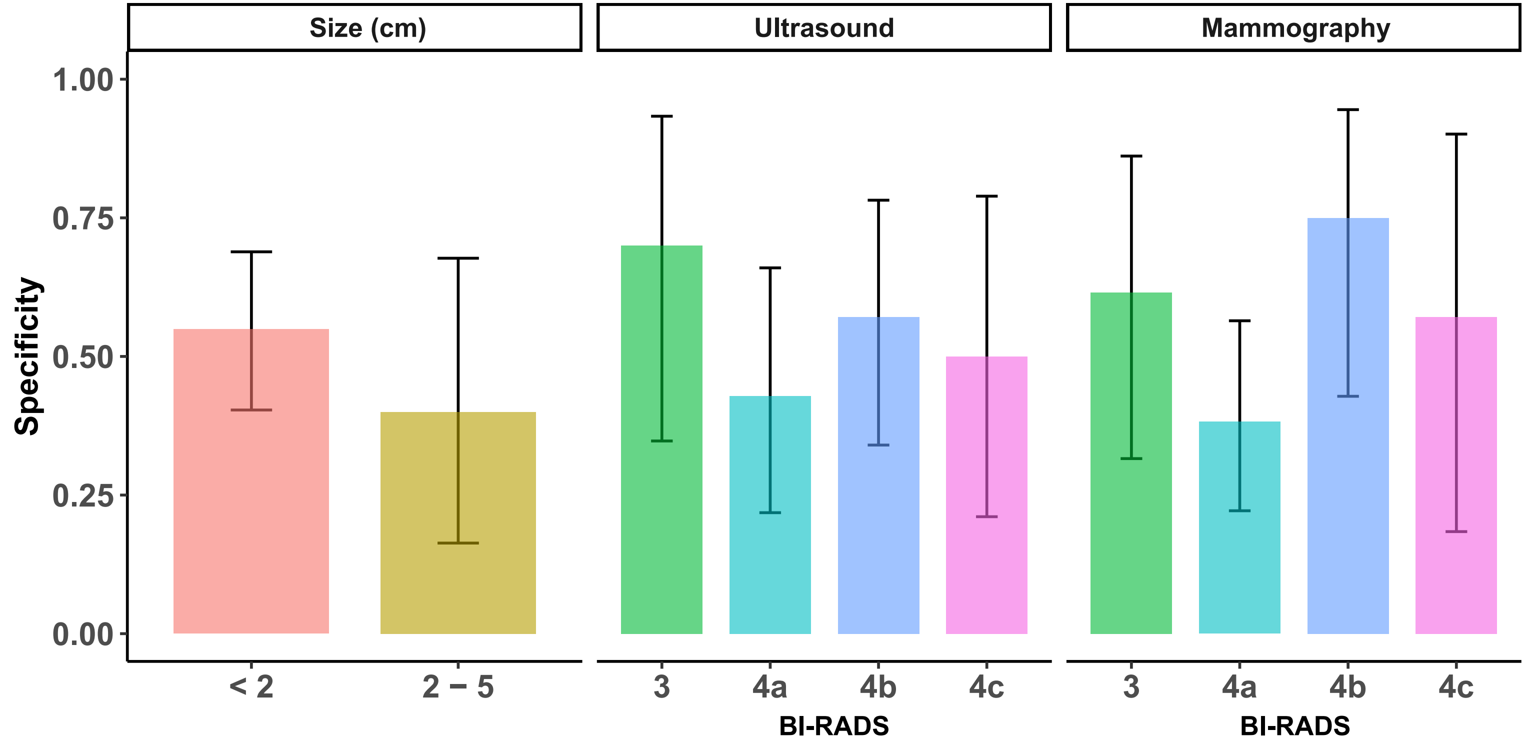

### Figure S11

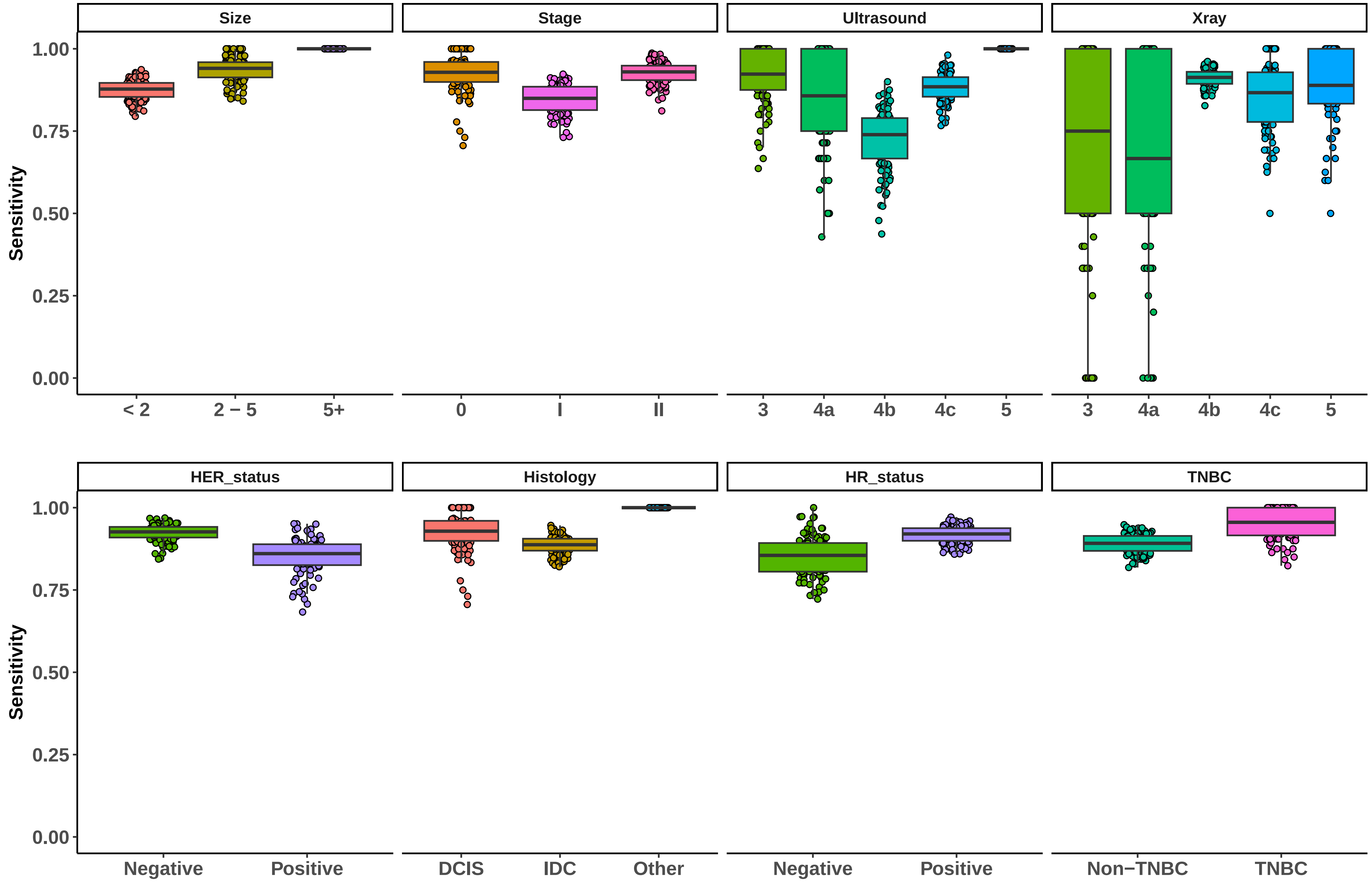

### Figure S12

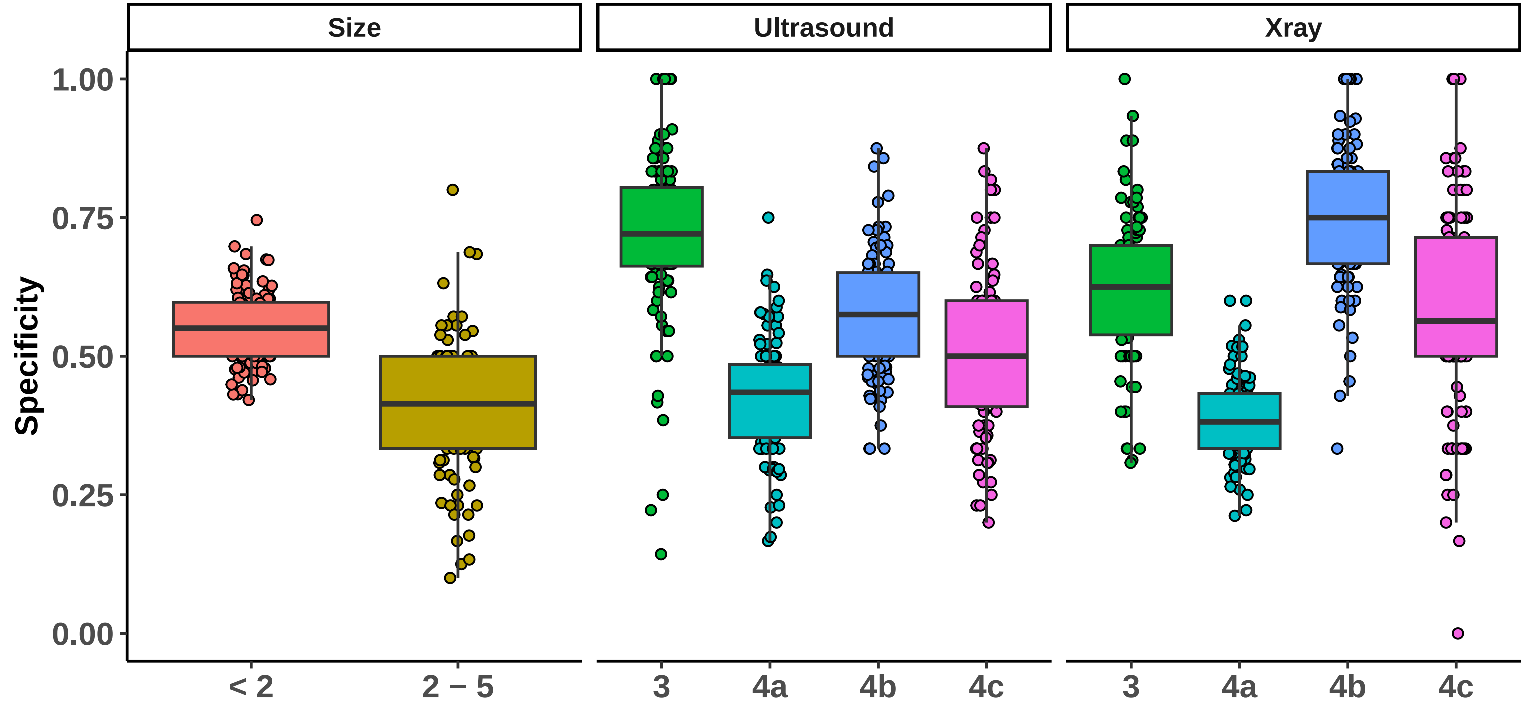

### Figure S13

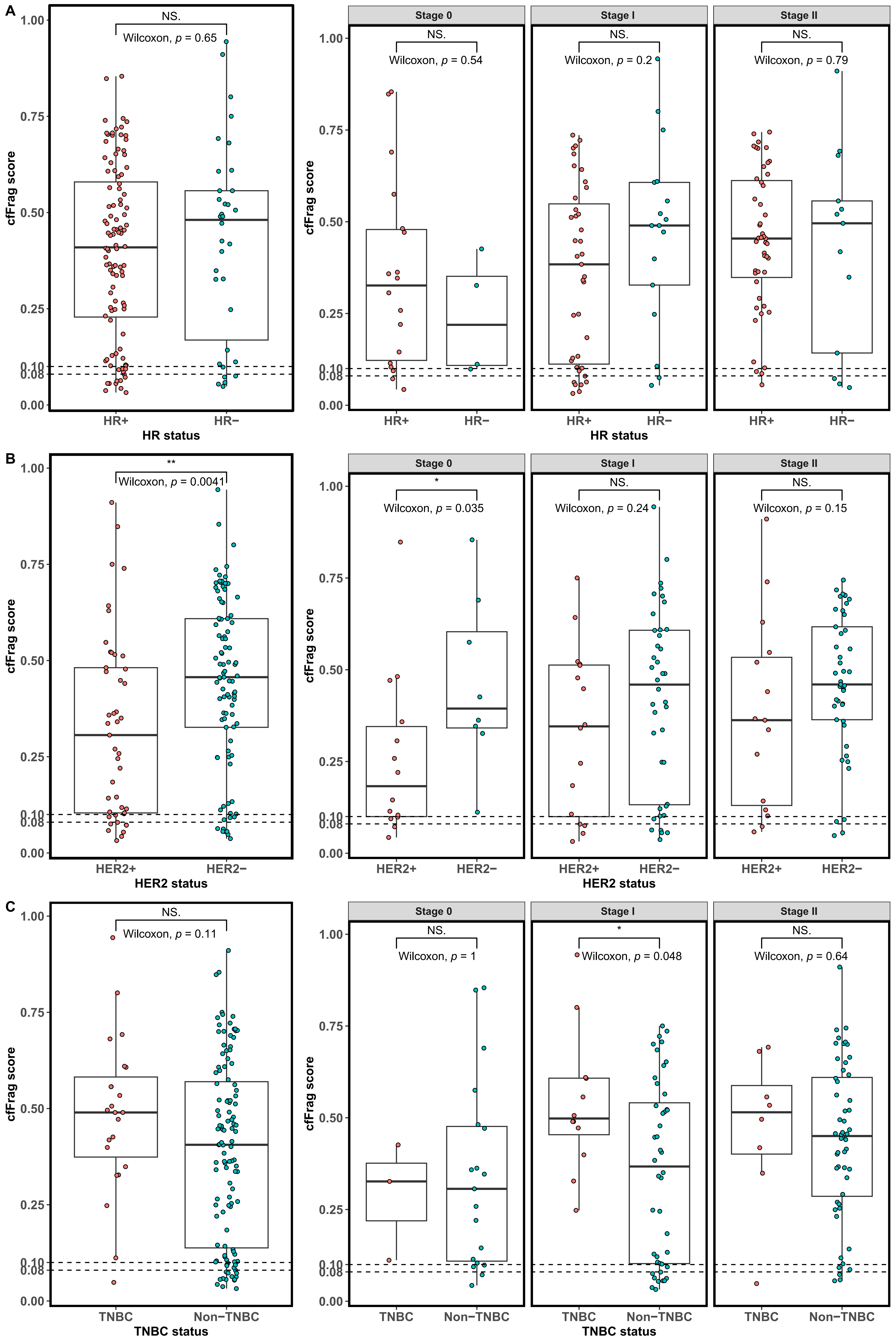

### Figure S14

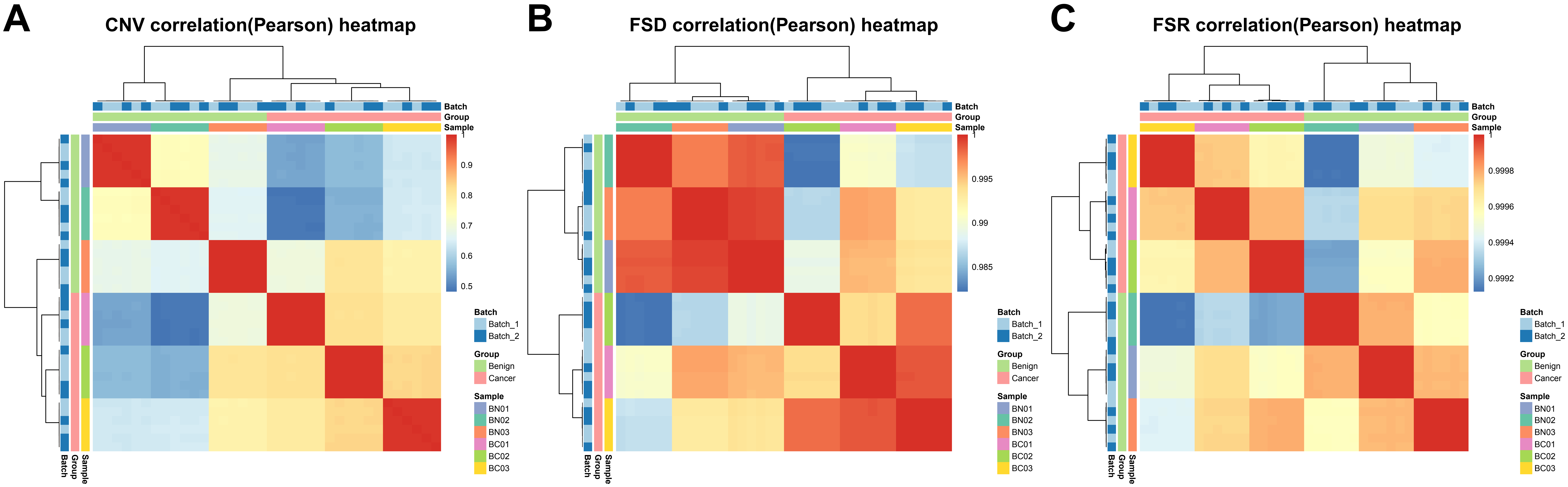

### Figure S15

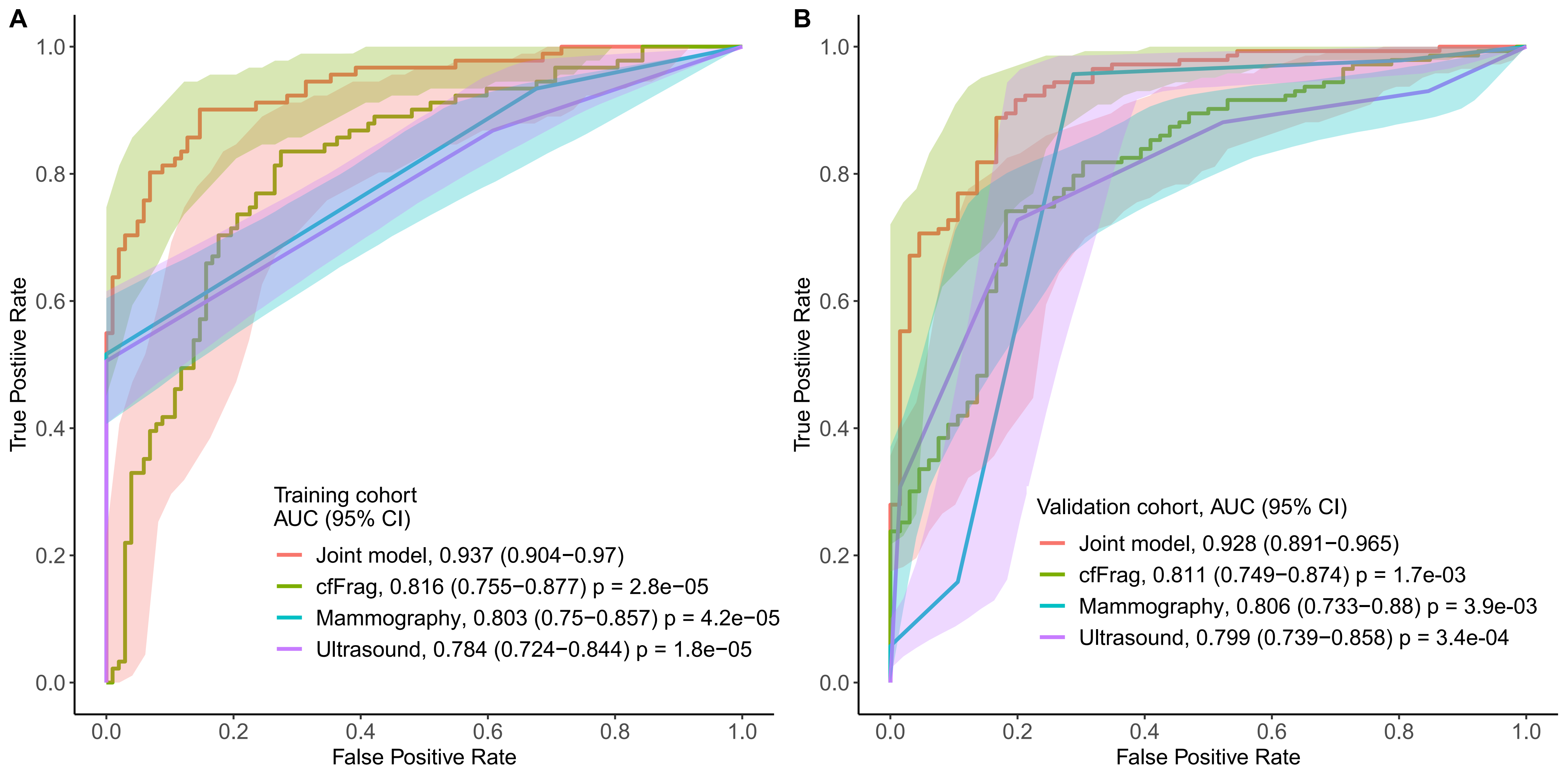
