## Supplementary material for "Cell-free DNA Fragmentomics Assay to Discriminate the Malignancy of Breast Nodules and Evaluate Treatment Response": Table S1

### Table S1. Patient Characteristics.

|  | **Training cohort**  **(Yantai cohort, N = 193)** | | | **Prospective validation cohort**  **(Beijing cohort, N = 209)** | |
| --- | --- | --- | --- | --- | --- |
|  | **Breast cancer**  **(N = 91)** | **Benign nodule**  **(N = 102)** | **Breast cancer**  **(N = 143)** | | **Benign nodule**  **(N = 66)** |
| **Age** | | | | | |
| Mean (SD) | 56.1 (8.9) | 40.6 (12.6) | 51.9 (11.9) | | 45.5 (12.0) |
| Median [Min, Max] | 57 [34-80] | 38 [18-90] | 52 [24-79] | | 46 [24-72] |
| **Stage** ^a,b^ | | | | | |
| 0/DCIS | 8 (8.8%) | - | 23 (16.0%) | | - |
| I | 33 (36.3%) | - | 57 (39.9%) | | - |
| II | 50 (54.9%) | - | 60 (42.0%) | | - |
| III-IV | 0 (0%) | - | 0 (0%) | | - |
| Unknown | - | - | 3 (2.1%) | | - |
| **Histology** ^a^ | | | | | |
| IDC | 78 (85.7%) | - | 115 (80.4%) | | - |
| DCIS | 8 (8.8%) | - | 23 (16.0%) | | - |
| Other | 5 (5.5%) | - | 5 (3.5) | | - |
| **Tumor size** ^a^ | | | | | |
| < 2 cm | 34 (37.4%) | 83 (81.4%) | 92 (64.3%) | | 51 (77.3%) |
| 2-5 cm | 52 (57.1%) | 19 (18.6%) | 50 (35.0) | | 15 (22.7%) |
| > 5 cm | 5 (5.5%) | - | 1 (0.7%) | | - |
| **HR status** ^a^ | | | | | |
| Positive | 63 (69.2%) | - | 105 (73.4%) | | - |
| Negative | 27 (29.7%) | - | 35 (24.5%) | | - |
| Unknown | 1 (1.1%) | - | 3 (2.1%) | | - |
| **HER2 status** ^a^ | | | | | |
| Positive | 19 (20.9%) | - | 46 (32.2%) | | - |
| Negative | 71 (78.0%) | - | 94 (65.7%) | | - |
| Unknown | 1 (1.1%) | - | 3 (2.1%) | | - |
| **TNBC status** ^a^ | | | | | |
| TNBC | 17 (18.7%) | - | 23 (16.1%) | | - |
| Non-TNBC | 73 (80.2%) | - | 117 (81.8%) | | - |
| Unknown | 1 (1.1%) | - | 3 (2.1%) | | - |
| **Mammography BI-RADS** ^a^ | | | | | |
| 3 | - | - | 3 (2.1%) | | 13 (19.7%) |
| 4a | 6 (6.6%) | 33 (32.4%) | 3 (2.1%) | | 34 (51.5%) |
| 4b | 38 (41.8%) | 69 (67.6%) | 111 (77.6%) | | 12 (18.2%) |
| 4c | 28 (30.8%) | - | 14 (9.8%) | | 7 (10.6%) |
| 5 | 19 (20.8%) | - | 8 (5.6%) | | - |
| NA | - | - | 4 (2.8%) | | - |
| **Ultrasound BI-RADS** ^a^ | | | | | |
| 3 | - | - | 10 (7.0%) | | 10 (15.2%) |
| 4a | 12 (13.2%) | 40 (39.2%) | 7 (4.9%) | | 21 (31.8%) |
| 4b | 33 (36.2%) | 62 (60.8%) | 22 (15.4%) | | 21 (31.8%) |
| 4c | 28 (30.8%) | - | 60 (41.9%) | | 12 (18.2%) |
| 5 | 18 (19.8%) | - | 44 (30.8%) | | 1 (1.5%) |
| NA | - | - | - | | 1 (1.5%) |

^a^ Data shown as the number (percentage).

^b^ Clinical staging was determined according to the eighth edition of the classification for breast cancer of the American Joint Commission of Cancer.

Abbreviation: N, number; SD, standard deviation; DCIS, ductal carcinoma *in situ*; IDC, invasive ductal carcinoma; HR, hormone receptor; HER2, human epidermal growth factor receptor 2; TNBC, triple-negative breast cancer; BI-RADS, Breast Imaging Reporting and Data System.
