## Supplementary material for "Cell-free DNA Fragmentomics Assay to Discriminate the Malignancy of Breast Nodules and Evaluate Treatment Response": Table S2

### Table S2. Selected top-performing base learners for constructing the final cfFrag model.

| Feature | Algorithm | Train AUC | Rank |
| --- | --- | --- | --- |
| CNV | DeepLearning | 0.791 | 1 |
|  | DeepLearning | 0.789 | 2 |
|  | GBM | 0.787 | 3 |
|  | GBM | 0.786 | 4 |
|  | DeepLearning | 0.739 | 5 |
|  | DeepLearning | 0.716 | 6 |
|  | XGBoost | 0.663 | 7 |
|  | XGBoost | 0.661 | 8 |
| FSR | DeepLearning | 0.754 | 1 |
|  | DeepLearning | 0.750 | 2 |
|  | DeepLearning | 0.750 | 3 |
|  | DeepLearning | 0.736 | 4 |
|  | DeepLearning | 0.691 | 5 |
|  | GBM | 0.667 | 6 |
|  | GBM | 0.654 | 7 |
|  | DeepLearning | 0.647 | 8 |
| FSD | DeepLearning | 0.750 | 1 |
|  | DeepLearning | 0.747 | 2 |
|  | DeepLearning | 0.744 | 3 |
|  | DeepLearning | 0.744 | 4 |
|  | DeepLearning | 0.703 | 5 |
|  | DeepLearning | 0.677 | 6 |
|  | DeepLearning | 0.655 | 7 |
|  | GBM | 0.631 | 8 |

Abbreviation: AUC, area under the curve; GBM, gradient boosting machine; CNV, copy number variation; FSD, fragment size distribution; FSR, fragment size ratio.
