## Supplementary material for "Cell-free DNA Fragmentomics Assay to Discriminate the Malignancy of Breast Nodules and Evaluate Treatment Response": Table S6

### Table S6. Evaluating the Fragmentomics Model Performances at 85% Sensitivity Cutoff.

| Training cohort  (5-fold cross-validation) | | **Actual** | | Prospective  validation cohort | | Actual | |
| --- | --- | --- | --- | --- | --- | --- | --- |
|  |  | BCs | BNs |  |  | BCs | BNs |
| **Predict** | BCs | 77 | 35 | **Predict** | BCs | 118 | 26 |
|  | BNs | 14 | 67 |  | BNs | 25 | 40 |
| Sensitivity (95% CI) | | 84.6% (75.5-91.3%) | | Sensitivity (95% CI) | | 82.5% (75.3-88.4%) | |
| Specificity (95% CI) | | 65.7% (55.6-74.8%) | | Specificity (95% CI) | | 60.6% (47.8-72.4%) | |
| PPV (95% CI) | | 68.8% (59.3-77.2%) | | PPV (95% CI) | | 81.9% (74.7-87.9%) | |
| NPV (95% CI) | | 82.7% (72.7-90.2%) | | NPV (95% CI) | | 61.5% (48.6-73.3%) | |
| Accuracy (95% CI) | | 74.6% (67.9-80.6%) | | Accuracy (95% CI) | | 75.6% (69.2-81.3%) | |

Abbreviation: BC, breast cancer; BN, benign nodule; CI, confidence interval; PPV, positive predictive value; NPV, negative predictive value.
